# Body composition subphenotypes, cardiometabolic risk and incident outcomes: validation in the population-based NAKO and UK Biobank imaging cohorts

**DOI:** 10.64898/2026.06.18.26355957

**Authors:** Elena Grune, Tobias Haueise, Marc-Nicolas von Itter, Matthias Jung, Fabian Bamberg, Saima Bibi, Christoph M. Friedrich, Patricia Fromherz, Hans-Ulrich Kauczor, Elias Kellner, Anna Köttgen, Lilian Krist, Thomas Kröncke, Wolfgang Lieb, Jürgen Machann, Johanna Nattenmüller, Fiona Niedermayer, Thoralf Niendorf, Tobias Nonnenmacher, Tobias Norajitra, Tobias Pischon, Marco Reisert, Christopher L. Schlett, Jeanette Schulz-Menger, Jakob Weiß, Annette Peters, Anne-Laure Boulesteix, Susanne Rospleszcz

## Abstract

**Background:** Anthropometric measures do not adequately capture heterogeneity in body fat distribution and corresponding cardiometabolic risk, whereas magnetic resonance imaging (MRI) enables precise differentiation and quantification of adipose tissue compartments and ectopic fat. We aimed to validate previously derived MRI-based body composition subphenotypes and their cardiometabolic risk profiles in two independent European cohorts.

**Methods:** Using deep learning–based image analysis, we quantified bone marrow, visceral, subcutaneous, cardiac, renal sinus, hepatic, skeletal muscle, and pancreatic fat in the imaging substudies of two population-based cohorts: the German National Cohort (NAKO, N=29,314, age range 19-74 years) and the UK Biobank (N=36,109, age range 40-69 years). Body composition subphenotypes, previously identified by k-means clustering, were evaluated using a rigorous statistical cluster validation framework with method-based and results-based approaches. In NAKO, cross-sectional associations between subphenotypes and estimated cardiovascular disease risk scores were examined using linear regression. In UK Biobank, longitudinal associations between subphenotypes and incident cardiometabolic outcomes, ascertained through hospital record linkage, were analysed using Cox regression.

**Findings:** All five body composition subphenotypes were robustly validated across both cohorts, and showed distinct fat distribution patterns and cardiometabolic risk profiles: I “lean”, II “average adiposity”, III “bone and muscle adiposity”, IV “hepato-abdominal adiposity”, and V “general and pancreatic adiposity”. Subphenotypes I–III showed progressive adipose tissue remodelling patterns likely reflecting ageing trajectories. The “hepato-abdominal adiposity” subphenotype showed highest risk of incident diabetes, whereas the “general and pancreatic adiposity” subphenotype showed highest overall cardiovascular disease burden and metabolic impairment.

**Interpretation:** MRI-derived body composition subphenotypes represent distinct fat distribution patterns that reflect ageing- and disease-related processes, which supports the potential of body composition phenotyping for improved cardiometabolic risk stratification and targeted prevention.

## Introduction

The importance of adipose tissue distribution beyond body mass index (BMI) in cardiometabolic risk is increasingly recognized [1, 2]. While the metabolic roles of visceral adipose tissue (VAT) and subcutaneous adipose tissue (SAT) have long been characterized [3], growing attention is turning to ectopic fat accumulation and the distinct local and systemic effects of individual fat depots [4]. Among these, liver fat has been bidirectionally linked to type 2 diabetes, with shared metabolic and endocrinologic pathways driving the progression of each condition [5]. Previous studies on adipose tissue distribution derived by magnetic resonance imaging (MRI) have jointly assessed abdominal, hepatic, and muscular fat to link their distribution to coronary heart disease, type 2 diabetes, or elevated metabolic risk [6, 7]. These findings highlight the importance of a multivariable approach for understanding the relationship between excess adiposity and cardiometabolic health, which provides more insight than anthropometric measures or single fat depots alone.

Using population-based MRI data, we previously identified five distinct body composition subphenotypes characterized by specific adipose tissue distribution patterns [8]. These subphenotypes reflected different cardiometabolic risk profiles beyond total adiposity [8]. We now aimed to 1) validate the subphenotypes using MRI data from the population-based German National Cohort (NAKO) and UK Biobank studies, and 2) to evaluate their associations with cardiovascular disease (CVD) risk and incident cardiometabolic outcomes.

## Methods

### Study populations

We used data from two deeply phenotyped population-based cohorts: NAKO (N = 205,415 [9]) and UK Biobank (N = 502,000 [10]).

In the NAKO imaging substudy, 30,868 individuals aged 19–74 years were examined between 2014 and 2019. Imaging was conducted within 12 weeks of the baseline assessment in participants without contraindications [11]. Whole-body MRI scans were acquired on 3 Tesla scanners (MAGNETOM Skyra, Siemens Healthineers, Erlangen, Germany) at five MRI centers with identical software and hardware settings. The examination programm covered the cerebrospinal, musculoskeletal, cardiovascular and thoracoabdominal system [11]. Image quality was assessed by trained radiologists and automatically derived quality measures [12].

The UK Biobank recruited participants aged 40-69 years between 2006 and 2010. In 2014, the imaging enhancement started with 100,000 participants re-invited for multi-modal imaging until 2025. We accessed all 36,515 image data sets available through May 2023. Participants without contraindications underwent abdominal and cardiac MRI at four dedicated assessment centres in England on 1.5 Tesla scanners (MAGNETOM Aera, Siemens Healthineers, Erlangen, Germany) with harmonised software and hardware. Image quality was ensured through standardised radiographer training and visual inspection during and after scanning [13].

Using data from T1-weighted 3D VIBE Dixon imaging acquired in axial orientation in both studies, we derived body composition measures including bone marrow, heart, liver, pancreas, kidney sinus, and skeletal muscle fat fractions (%), as well as VAT, SAT (L), and intramuscular adipose tissue (dm^3^) volumes. Segmentation and fat quantification were performed fully automatically using validated deep-learning-based algorithms in both cohorts, ensuring consistent measurements (Supp Text 1).

### Ethics

Both studies were performed with the approval of the relevant local ethics committees and in accordance with national law and the Declaration of Helsinki. All participants gave written informed consent before the examinations.

### Outcome and Covariate Assessment

In NAKO (no incident outcome data after MRI exam available), short- and long-term CVD risk was estimated by SCORE2, 10-year Framingham Risk Score (FRS10), and 30-year Framingham Risk Score (FRS30, Supp Text 2).

In UK Biobank (no risk score data at MRI exam available), incident outcomes included diabetes, myocardial infarction, ischemic stroke, major adverse cardiovascular events (MACE; defined as myocardial infarction or ischemic stroke or mortality from major CVD), and hypertension. Outcomes were ascertained using ICD-9 and ICD-10 codes based on linkage to hospital inpatient records. All-cause mortality was obtained through linkage to national death registries. Outcome time horizons differed between cohorts, with a median follow-up of approximately 4 years in UK Biobank versus 10- and 30-year estimates in NAKO. Details on outcome and covariate definitions are provided in Supp Text 2.

### Validation of subphenotpyes

We validated the previously identified body composition subphenotypes by adopting the cluster validation framework developed by Ullmann et al. [14], using both a method-based (NAKO) and a result-based (UK Biobank) validation approach. Method-based validation aims to assess if re-applying the clustering method to new data identifies similar structures, while result-based validation assesses whether the existing clustering remains meaningful when transferred to new data [14]. Method-based validation in NAKO corresponded to a de novo clustering, where the same preprocessing (sex-specific standardization), clustering algorithm (Hartigan-Wong k-means) and number of clusters (k = 5) were used as in the original study [8]. Clusters, corresponding to body composition subphenotypes, were labeled with Roman numerals I to V, sorted by ascending average VAT [8]. Cluster stability was assessed by a similarity measure (Jaccard index) across repeated clustering on 1000 bootstrap samples. Internal (relative cluster size, adipose tissue distribution) and visual cluster properties (principal component biplots, silhouette plots) were compared with the original subphenotypes [8]. As sensitivity analysis, clustering was performed separately for women and men in NAKO.

Result-based validation in the UK Biobank assigned participants to NAKO-derived clusters using a nearest-centroid classifier (Euclidean distance), thereby transferring individuals to the closest cluster instead of re-running the clustering algorithm. Prior to cluster transfer, adipose tissue measures were standardized using two complementary approaches to address different aspects of cross-cohort comparability. First, UK Biobank data were sex-specifically standardized internally (mean = 0, standard deviation = 1 within UK Biobank) to assess whether adipose tissue distribution patterns corresponding to the NAKO-derived clusters are robust to differences in adipose tissue measurements and population characteristics. Second, UK Biobank data were sex-specifically standardized using the means and standard deviations from NAKO to assess transferability of the cluster framework on the original NAKO scale and the impact of cohort specific characteristics on cluster allocation. To assess external cluster properties based on variables not included in clustering, associations with CVD risk scores and incident outcomes were compared.

### Statistical analysis

Adipose tissue data, risk factors and risk scores are given as mean and standard deviation or median and interquartile range for continuous variables and as count and percentage for categorical variables.

Adipose tissue distribution within subphenotypes of body composition is graphically illustrated by radar charts. Association of subphenotypes with log-transformed CVD risk scores in NAKO was assessed by linear regression and estimates are reported as exponentiated beta coefficients, corresponding to percent change in geometric mean.

We performed additional descriptive analyses within clinically defined risk subgroups in NAKO. First, we examined a subgroup with increased CVD risk as defined by both elevated BMI (25–30 kg/m^2^) and waist circumference (≥90 cm in women, ≥100 cm in men), and summarized adipose tissue patterns across subphenotypes within this group. Second, we evaluated subphenotype-specific CVD risk across clinically relevant strata, calculating median estimated CVD risk within clusters by age groups, hypertension status, BMI categories, and diabetes status.

Associations between subphenotypes and incident outcomes in UK Biobank were assessed with cumulative incidence curves and Cox proportional hazards regression models, adjusted for 1) age and sex, 2) additionally for BMI, smoking status, and alcohol consumption. The proportional hazards assumption was tested by assessing time-dependence of Schoenfeld residuals. Results were reported as hazard ratio (HR) with 95% confidence intervals (CI).

P-values <0.05 were considered to indicate statistical significance. All analyses were conducted with R (version 4.3.3).

## Results

### Study samples

The final NAKO sample comprised 29,314 individuals (mean age 48.2±12.3 years, 44.3% women) and the final UK Biobank sample 36,109 individuals (65.0±7.8 years, 51.7% women) with complete adipose tissue data (Supp Table 1).

Adipose tissue depots and cardiometabolic risk markers showed distinct distributions across the NAKO study population (Supp Tables 2). Adipose tissue distribution differed by sex: men exhibited higher levels of bone marrow fat fraction (BMFF), VAT, cardiac fat fraction (CFF), renal sinus fat fraction (RFF), hepatic fat fraction (HFF), and pancreatic fat fraction (PFF), whereas women had higher SAT, skeletal muscle fat fraction (SMFF), and intramuscular adipose tissue (IMAT). The median 10-year CVD risk, as estimated by SCORE2, was 4.8% in men and 2.2% in women. Estimates based on FRS10 and FRS30 showed a comparable distribution (Supp Table 2).

In UK Biobank, adipose tissue showed largely similar sex-specific distributions, but higher BMFF in women. During follow-up (median 4.2 years), 613 incident MACE, 584 incident diabetes, and 2,667 hypertension cases occurred. A total of 625 deaths were recorded over a median follow-up of 4.8 years (Supp Table 2).

### Cluster validation

We conclude robust validation of the five subphenotypes in both cohorts based on adipose tissue distribution patterns and structures consistent with the original Clusters: Cluster I (“Lean”), Cluster II (“Average adiposity”), Cluster III (“Bone and muscle adiposity”), Cluster IV (“Hepato-abdominal adiposity”), and Cluster V (“General and pancreatic adiposity”). Both UK Biobank standardization approaches in result-based validation yielded highly concordant fat distribution patterns across clusters, supporting the robustness and cross-cohort transferability of the identified subphenotypes. In the following, we present results for cluster assignment using NAKO-based standardization.

Relative cluster sizes were similar in both cohorts, with Cluster II being the largest and Clusters IV and V the smallest (Supp Figure 1). Relative adipose tissue distributions within subphenotypes were consistent across NAKO, UK Biobank and the original study (Supp Figure 2). Cluster stability was high for Clusters I-III and V with Jaccard Indices >0.75, and moderate for Cluster IV (Supp Table 3). Principal component analysis and silhouette plots indicated consistent cluster separation and cohesion in NAKO, UK Biobank and the original study (Supp Figure 3 and 4). Relative CVD risk by cluster largely followed the same pattern as in the original study. Compared to Cluster I, all clusters showed notably increased risk, with Cluster V showing the highest increase (Supp Figure 5).

In sex-stratified clustering as sensitivity analysis, adipose tissue distribution patterns were largely consistent with the main analysis. In women, Cluster IV was more strongly characterized by abdominal fat, while hepatic fat was not elevated. Instead, hepatic fat was highest in Cluster V (Supp Figure 6). Sex-specific CVD risk patterns aligned with these distributional differences: in women, Cluster IV was less strongly associated with increased risk than in the main analysis, while Cluster V remained the cluster with the highest risk elevation in both analyses (Supp Figure 7).

### Body composition subphenotypes and cardiovascular disease risk in NAKO

Body composition subphenotypes are described by radar charts depicting the average adipose tissue distribution profiles in each cluster compared to the average of the whole sample (Figure 1). “Lean” subphenotype I had lowest average age (36.5 years) and lowest prevalence of metabolic and lifestyle risk factors (Supp Table 4). Correspondingly, short- and long-term cardiovascular risk was lowest across all subphenotypes (median SCORE2 1.7%, median FRS10 3.7%, Table 1). “Average adiposity” subphenotype II had a mean age of 47.3 years, diabetes prevalence of 2.0%, and hypertension prevalence of 28.9% (Supp Table 4). Cardiovascular risk was 1.6–2.3-fold higher compared with the lean subphenotype I (Figure 2). “Bone and muscle adiposity” subphenotype III had a mean age of 55.2 years and 75.2% of women were post menopause (Table 1). Cardiovascular risk was 2.7–3.5-fold higher compared to subphenotype I. “Hepato-abdominal adiposity” subphenotype IV had a mean age of 51.4 years, highest proportion of diabetes (15.9%) of all subphenotypes and highest liver enzyme levels (Supp Table 4). Cardiovascular risk was 2.9–3.8-fold higher compared to subphenotype I. “General and pancreatic adiposity” subphenotype V had highest average age (59.4 years), with 86.3% of women being post menopause. Although average BMI was comparable to that of subphenotype IV (32.2 kg/m^2^, Table 1), short- and long-term cardiovascular risk was substantially higher (Figure 2).

In the subgroup of 1,557 women and 2,199 men with elevated BMI and waist circumference, reflecting current guidelines to combine anthropometric measures to evaluate excess adiposity [1], all five subphenotypes were represented (Supp Figure 8). This subset of individuals showed little variation in SAT across subphenotypes, while distribution patterns of the other adipose tissues corresponded to the those in the overall sample. In the subgroups of individuals with diabetes or hypertension, substantially lower short- and long-term CVD risk was observed if their body composition corresponded to subphenotype I or II (Figure 3). Subphenotypes provided stronger short- and long-term risk stratification than BMI categories (Figure 3).

**Table 1.**
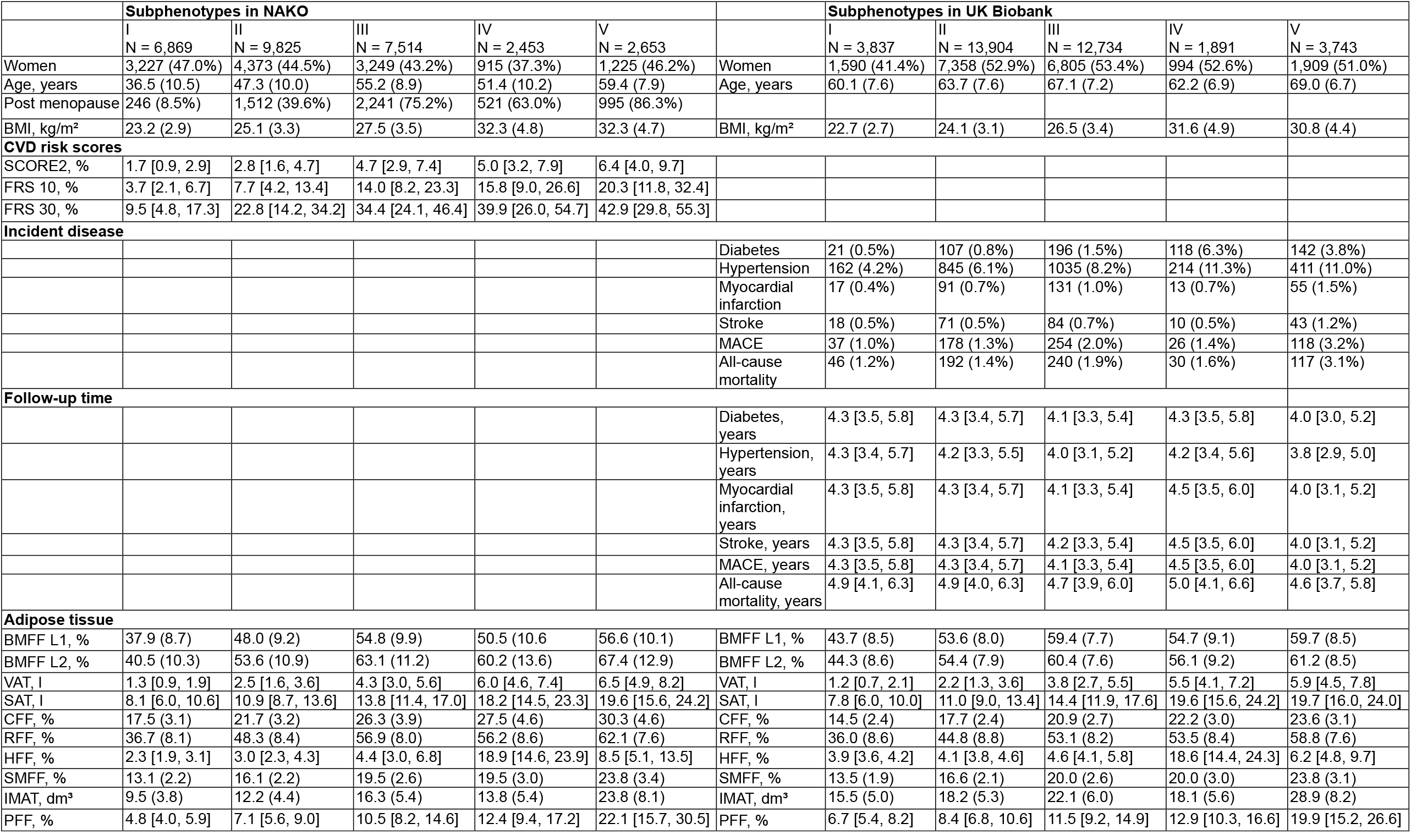
Characteristics of the five body composition subphenotypes in NAKO and UK Biobank. Values are given as mean (standard deviation) or median [interquartile range] for continuous values and counts (percentage) for categorical values. Abbreviations. BMI: Body mass index, SCORE2: Systematic Coronary Risk Evaluation Model 2, FRS: Framingham Risk Score. MACE: major adverse cardiovascular events, BMFF L1/L2: bone marrow fat fraction at vertebrae L1/L2, VAT: visceral adipose tissue, SAT: subcutaneous adipose tissue, CFF: Cardiac fat fraction, RFF: renal sinus fat fraction, HFF: hepatic fat fraction, SMFF: skeletal muscle fat fraction, IMAT: intramuscular adipose tissue, PFF: pancreatic fat fraction.

**Figure 1.**
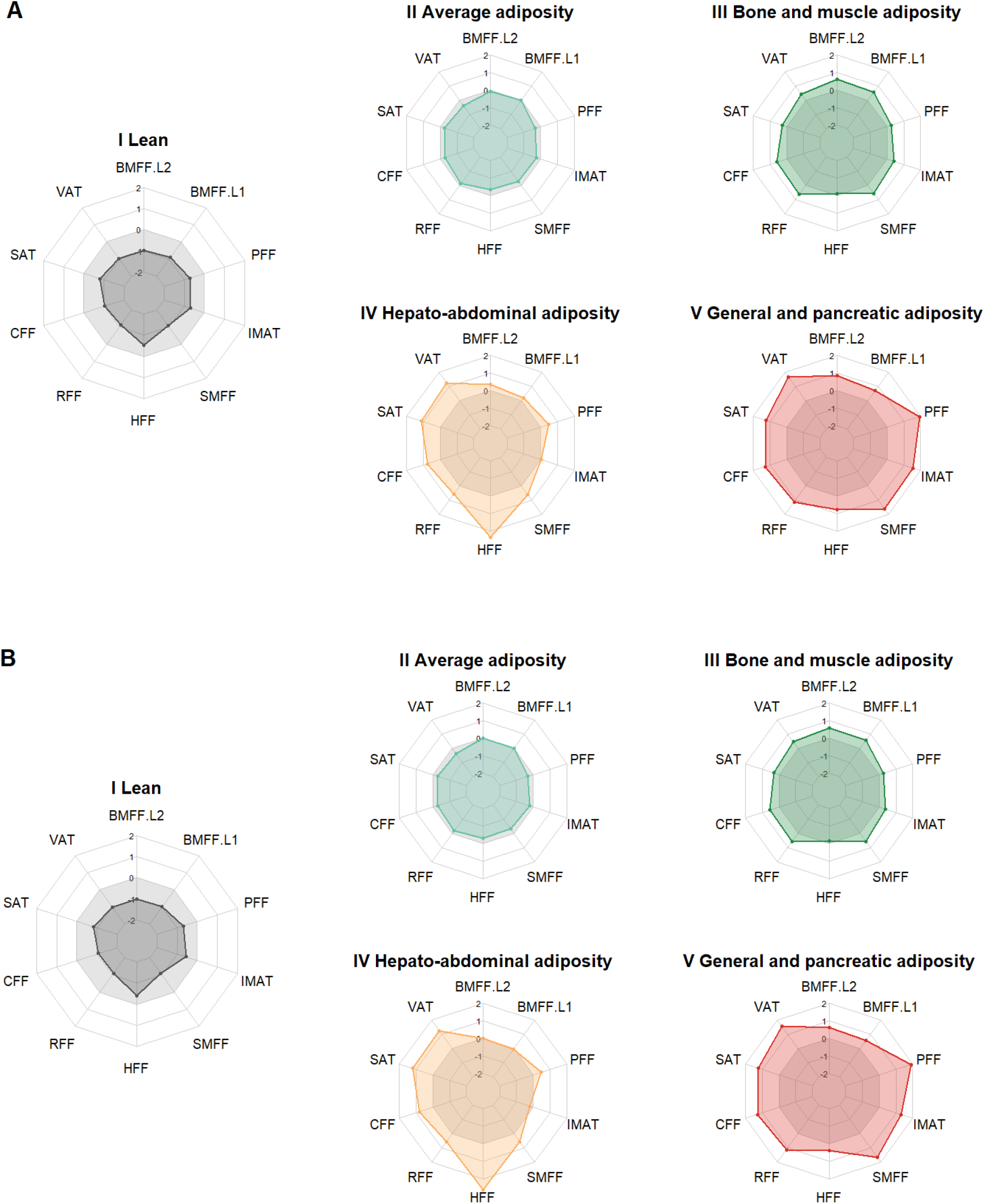
The five body composition subphenotypes in NAKO and UK Biobank. Radar charts of standardized adipose tissue depots (z-scores) across the five Clusters, representing body composition subphenotypes in NAKO (panel A) and UK Biobank (panel B). The respective population means are shown as grey polygon at 0. Colored lines represent mean values within each cluster. Axes range from - 2 to +2, corresponding to standard deviation from the mean. Abbreviations. BMFF L1/L2: bone marrow fat fraction at vertebrae L1/L2, VAT: visceral adipose tissue, SAT: subcutaneous adipose tissue, CFF: cardiac fat fraction, RFF: renal sinus fat fraction, HFF: hepatic fat fraction, SMFF: skeletal muscle fat fraction, IMAT: intramuscular adipose tissue, PFF: pancreatic fat fraction.

**Figure 2.**
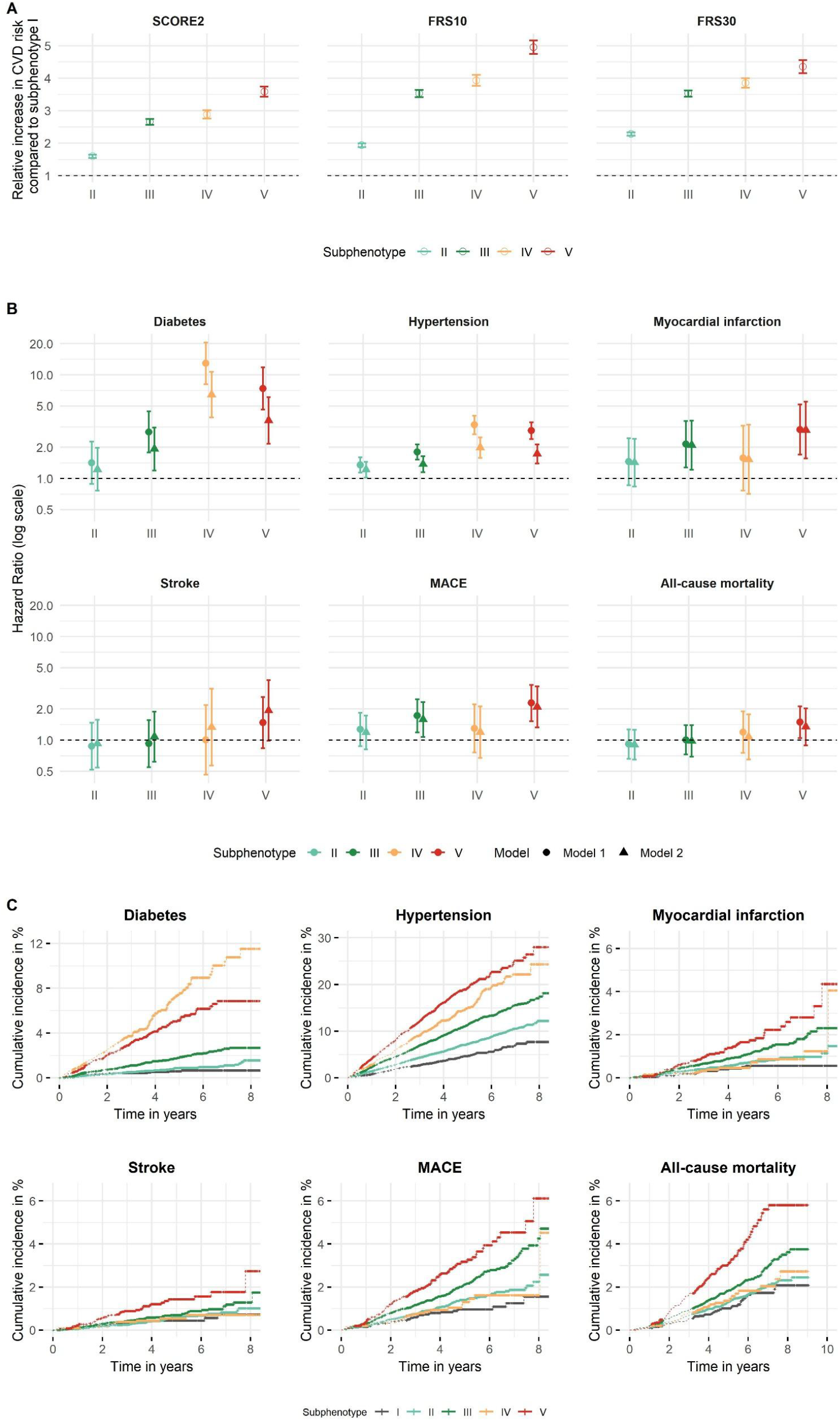
Association of body composition subphenotypes with CVD risk in NAKO and cardiometabolic outcomes in UK Biobank Panel A. Effect estimates of the association between subphenotypes and estimated CVD risk, derived from linear regression models using the log-transformed SCORE2, FRS10, FRS30 risk score, respectively, as outcome. Estimates are shown as exponentiated beta coefficients with 95% CI, representing the percentage change in the geometric mean. Since risk score calculation included age and sex, the regression models were unadjusted. Subphenotype I served as reference category. Panel B. Hazard ratios from Cox proportional hazards regression models with 95% confidence intervals. Models were adjusted for (1) age and sex and (2) additionally for BMI, smoking status, and alcohol consumption. Subphenotype I served as reference category. Panel C. Cumulative incidence curves for cardiometabolic outcomes and all-cause mortality across subphenotypes. Abbreviations. CVD: cardiovascular disease, FRS: Framingham Risk Score, MACE: major adverse cardiovascular events.

**Figure 3.**
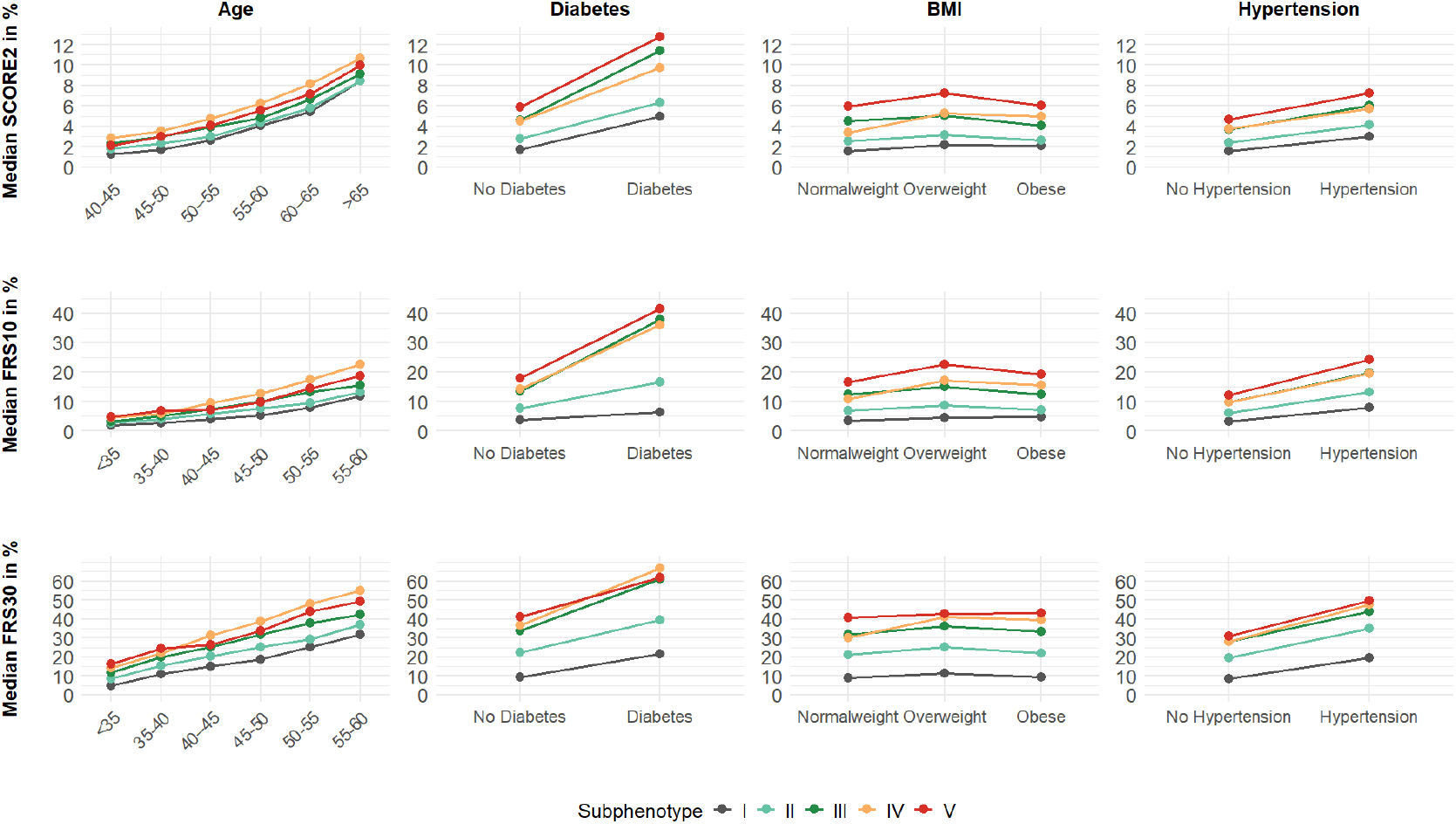
Subphenotype specific CVD risk across clinical subgroups in NAKO. Median estimated CVD risk is shown for each subphenotype stratified by age categories, diabetes status, BMI categories, and hypertension status. Abbreviations. FRS: Framingham Risk Score.

### Body composition subphenotypes and incident cardiometabolic disease in UK Biobank

Relative distributions of age, BMI, and risk factors were comparable across subphenotypes in UK Biobank and NAKO (Supp Table 4). After follow-up, “lean” subphenotype I showed lowest cumulative incidence of MACE, MI, stroke and diabetes, and lowest overall mortality (Figure 2). “Average adiposity” subphenotype II showed higher cumulative incidence of these outcomes than subphenotype I, but hazards were not statistically significant (Figure 2). “Bone and muscle adiposity” subphenotype III showed significantly higher hazards of MI, MACE, hypertension and diabetes compared to subphenotype I (Figure 2). “Hepato-abdominal adiposity” subphenotype IV showed the highest cumulative incidence of diabetes across subphenotypes, with the highest hazards (HR 6.5, 95%CI [3.9, 10.7]) compared to subphenotype I. “General and pancreatic adiposity” subphenotype V showed significantly higher hazards for MACE, MI, stroke and diabetes compared to subphenotype I (Figure 2). There was no significant association with mortality for any subphenotype.

## Discussion

Subphenotypes of body composition represent distinct distribution patterns of a broad panel of adipose tissue depots. We validated these subphenotypes and their risk factor and outcome associations in NAKO and UK Biobank, two large independent population-based cohorts with MRI data. Our results provide strong evidence that the subphenotypes capture true biological patterns of adipose tissue distribution in European populations, reflecting underlying aging and disease processes.

Limitations of BMI in assessing individual fat distribution and cardiometabolic risk are well recognised [1, 7]. In our study, BMI provided inferior CVD risk stratification compared to the subphenotypes, and additional BMI adjustment had little effect on longitudinal outcome associations. Importantly, the subgroup of individuals with visceral adiposity as defined by elevated BMI and waist circumference still showed substantial heterogeneity in fat distribution and corresponding CVD risk, illustrating the challenge to assess cardiometabolic status solely by anthropometric markers. In contrast, MRI enables detailed, non-invasive 3D body composition assessment, including measurements of ectopic fat depots [15]. Therefore, body composition profiling based on MRI data has received increasing attention, particularly on UK Biobank data [6, 16], but also on patient collectives [17]. However, external validation still remains an issue. To overcome this limitation, we have used a rigorous statistical cluster validation framework to show the robustness and transferability of body composition subphenotypes on independent data.

We hypothesize that subphenotypes I-III indicate a relatively healthy regular aging trajectory, whereas subphenotypes IV and V represent distinct, high cardiometabolic risk states. “Lean” subphenotype I represented a metabolically favourable pattern characterized by healthy lifestyle, low disease prevalence, and lowest cardiometabolic risk. Since this phenotype was identified across cohorts with different age distributions, its younger age relative to the overall population might reflect low biological, rather than chronological age. Emerging research shows that MRI-based biological age estimation at organ and tissue level is feasible and may have prognostic value for health outcomes [18]. Overall, the “lean” subphenotype may represent a desirable body composition profile serving as a benchmark for cardiometabolic health.

The “average adiposity” subphenotype II showed moderate levels of adipose tissue depots compared to the “lean” subphenotype, accompanied primarily by a moderately higher age rather than pronounced metabolic impairment. No significant associations of this subphenotype with incident cardiometabolic outcomes were observed. This pattern might reflect adipose tissue remodeling with age, e.g. redistribution of white adipose tissue, expansion of bone marrow fat and increased ectopic fat accumulation [19]. Subphenotype II can be considered a critical juncture from where adipose tissue trajectories can take a more favorable or unfavorable turn.

“Bone and muscle adiposity” subphenotype III represents the most advanced stage in the suggested aging trajectory, characterized by particularly high muscle and bone marrow fat. Myosteatosis, fatty infiltration of the muscle, contributes to impaired muscle quality [20], whereas high bone marrow fat has been associated with declining bone strength [21]. Factors associated with bone marrow adipogenesis such as sex hormone deficiency are observed to also contribute to myosteatosis, supporting the concept of shared pathways underlying musculoskeletal and vascular ageing [22]. Overall, characteristics of subphenotype III likely reflect physiological hallmarks of ageing rather than markedly adverse adiposity, and underline the critical importance of maintaining musculoskeletal health and preventing frailty in older adults to reduce the risk of sarcopenia, osteoporosis, and functional decline [23].

In contrast to the suggested ageing trajectories, the “hepato-abdominal adiposity” subphenotype IV emerged as a distinct, metabolically high-risk group at a relatively younger age. Characterized by high SAT, VAT, and hepatic fat with increased levels of liver enzymes, this subphenotype had the highest incidence of diabetes. Lifestyle factors might have partly contributed to this pattern, since we observed lowest levels of physical activity in this subphenotype; however alcohol consumption was not substantially elevated. The bidirectional relationship between hepatic steatosis and T2D, where liver fat promotes hepatic insulin resistance, and diabetes further accelerates progression of steatotic liver disease, is well established [5]. Intriguingly, subphenotype IV showed some discrepancies between women and men, reflecting the well-known sex differences in hepatic fat distribution and steatosis prevalence [24]. Sex differences are similarly observed in hepatic steatosis progression, with women being post menopause shifting towards higher risk of fibrosis and higher excess risk of CVD [25]. From a clinical perspective, this subphenotype may be particularly relevant for targeted prevention and intervention strategies. Transition from a healthy aging trajectory to this unfavorable adipose tissue distribution might be due to specific dietary patterns or other environmental factors which could provide actionable targets. On the other hand, it would be relevant to understand whether this subphenotype has a genetic basis, as genetic determinants of fat distribution can help explain the role of specific depots in cardiometabolic disease [26]. Regarding intervention, a recent trial has shown that novel dual GIP and GLP-1 receptor agonist therapy can reduce abdominal and hepatic fat, as measured by MRI, in T2D patients, while also improving glycaemic control and liver enzyme levels [27].

The “general and pancreatic adiposity” subphenotype V was characterised by overall increased adiposity in all adipose tissue depots. The high levels of pancreatic fat in this subphenotype reflect substantial metabolic impairment, such as high prevalence of diabetes and hypertension, low kidney function and inflammation [28]. The high prevalence, but lower incidence of T2D, as compared with the “hepato-abdominal adiposity” subphenotype, also supports that pancreatic fat accumulation might reflect an advanced stage of sustained systemic adiposity. Previous studies have reported partly conflicting findings regarding the relationship between pancreatic fat, insulin secretion, and T2D, suggesting that fatty pancreas may interact with individual genetic risk for T2D rather than acting as an isolated pathogenic factor [29]. Overall, the “general and pancreatic adiposity” subphenotype appears consistent with the recently updated definition of clinical obesity as a chronic systemic disease with established organ dysfunction potentially leading to severe complications [1]. We observed clear organ and metabolic impairment, together with the highest estimated CVD risk and highest hazards of incident MACE, indicating advanced disease. From a clinical perspective, preventing progression towards this subphenotype is imperative, particularly as remission of manifested organ damage may not always be possible.

Our results provide important insights into the age and population structure of adipose tissue distribution, and lay the groundwork for future research: First, to identify potential prevention opportunities and intervention targets, we need to understand how individuals transition between subphenotypes over time and what drives these trajectories. Repeated imaging in UK Biobank and NAKO will enable such individual-level analyses. A comprehensive approach incorporating not only imaging but also diet, physical activity, and genetic factors is needed to understand the trajectories. Second, since all studied cohorts mainly comprised individuals of European ancestry, validation in more diverse populations is needed. Third, scalable alternatives to MRI-based profiling are needed for clinical translation. Although opportunistic screening using routine imaging data and automated analysis may enable wider application, clinical practice will also require less resource-intensive approaches. Novel blood-based biomarkers could be identified through proteomics and metabolomics studies. Early metabolomics approaches have shown promising results for reflecting body composition [30].

In conclusion, MRI-derived body composition subphenotypes capture heterogeneity in adiposity at the population level beyond BMI. The subphenotypes reflect distinct patterns of ageing and disease. Their robust validation across two European cohorts supports their transferability and provides a basis for differentiated characterisation of cardiometabolic risk and future prevention targets.

## Data Availability

The datasets analyzed during the current study are not publicly available. Access to and use of NAKO data and biosamples can be obtained via an electronic application portal (https://transfer.nako.de). Access to UK Biobank data is obtained to all bona fide researchers upon submitting a health-related research proposal to the UK Biobank (https://www.ukbiobank.ac.uk). The main analysis code of the current study is provided as a supplementary file.

## Acknowledgements

We thank all participants who took part in the NAKO and UK Biobank study and the staff of these research initiatives. MJ was supported by the Berta-Ottenstein Program for Advanced Clinician Scientists, Faculty of Medicine, University of Freiburg.

## CRediT author statement

Conceptualization: SR; Investigation: EG, SR; Methodology: EG, ALB, SR; Formal analysis: EG; Resources: EG, TH, MnvI, MJ, FB, SB, CMF, PF, HUK, EK, AK, LK, TK, WL, JM, JN, FN, TN, TNon, TNor, TP, MR, CLS, JSM, JW, AP, ALB, SR; Data Curation: EG, TH, MJ, EK, TNon, TNor, MR, SR; Visualization: EG; Writing - original draft: EG, SR; Writing - review and editing: EG, TH, MnvI, MJ, FB, SB, CMF, PF, HUK, EK, AK, LK, TK, WL, JM, JN, FN, TN, TNon, TNor, TP, MR, CLS, JSM, JW, AP, ALB, SR.

