## Supplementary Material for "Body composition subphenotypes, cardiometabolic risk and incident outcomes: validation in the population-based NAKO and UK Biobank imaging cohorts"

Elena Grune et al.

### Supplementary Texts

#### Supplementary Text 1:

*Image analysis in NAKO*

A total of 30,868 NAKO MRI scans were available and processed as described below. This resulted in 29,314 participants with complete adipose tissue data. Missing adipose tissue data were due to poor image quality, image artefacts, or technical malfunctions and were assumed to be missing at random.

Bone marrow adipose tissue was assessed as bone marrow fat fraction in % at vertebrae L1 and L2. The segmentation of the images acquired with a T1-weighted 3D VIBE two-point Dixon sequence was performed automatically using a deep-learning based 3D U-Net model [1].

VAT volume in l was obtained using the T1-weighted 3D VIBE two-point Dixon sequence from hip to cardiac apex using a deep-learning based automatic U-Net segmentation model [2].

SAT volume in l was obtained in pelvis, abdomen and thorax using the T1-weighted 3D VIBE two-point Dixon sequence using the Deep Neural patchworks segmentation framework [3].

Skeletal muscle fat fraction was measured in % based on the T1-weighted 3D two-point VIBE Dixon sequence. It included all trunk (autochthonous and chest/abdominal wall), pelvic, and proximal thigh muscles within the deep peripheral fascia from the upper plate of the first thoracic vertebra to the femoral insertion of the adductor brevis muscle. The fat fraction was derived as mean intensity of fat images divided by the sum of fat and water images. Intramuscular adipose tissue was quantified in $dm^3$ limited to the autochthonous spinal musculature [4].

Liver fat was quantified as proton density fat fraction in % based on a multi-echo Dixon VIBE sequence with six echo times. Prior to the U-Net based segmentation, swap artifacts in the images were detected and repaired using an automated approach [5].

A fully automated deep learning pipeline was developed to quantify pancreas volume and fat fraction in % from T1-weighted 3D VIBE two-point Dixon. The model was trained on manually annotated scans and validated internally and externally [6].

Cardiac fat was quantified as fat fraction (%) from T1-weighted 3D VIBE two-point Dixon images. Heart segmentation was performed using the VIBESegmentator software [7], and the resulting cardiac fat fraction reflects fat content within the segmented myocardium and adjacent tissue.

For the assessment of renal sinus fat, images from T1-weighted 3D VIBE two-point Dixon sequences were segmented into cortex, medulla, and sinus using a hierarchical, multi-scale U-net [8]. Fat in renal sinus was quantified as fat fraction in %, calculated separately for each kidney and then averaged across both kidneys.

*Image analysis in UKB*

UKB imaging data were accessed on May 25, 2023 and 36,515 available scans were identified. After excluding 278 individuals with corrupt or incomplete images, 36,237 scans were processed as described below. Complete adipose tissue measurements were available in 36,109 individuals (99.5%; Supplementary Table 1).

Bone marrow adipose tissue was assessed as bone marrow fat fraction in % at vertebrae L1 and L2. The segmentation of the images was performed automatically using VIBESegmentator.

VAT and SAT volume in l was obtained in pelvis, abdomen and thorax using the T1-weighted 3D VIBE two-point Dixon sequence using the Deep Neural patchworks segmentation framework [3].

Skeletal muscle fat fraction, intramuscular adipose tissue, cardiac, pancreatic and renal sinus fat fraction was quantified as described for NAKO above.

**Supplementary Text 2:**

*Outcome and Covariate Assessment in NAKO*

CVD risk estimation was restricted to participants without a history of CVD and within the applicable age ranges for each model, in line with the populations used for their original derivation and validation. All risk scores included common core predictors: age, sex, systolic blood pressure, smoking status, total cholesterol, HDL cholesterol, and diabetes. Use of antihypertensive medication was additionally included in both Framingham models. Regarding endpoint definitions, SCORE2 estimates the 10-year risk of fatal CVD and non-fatal myocardial infarction and stroke, in individuals aged 40–69 years [9]. For participants with diabetes, the SCORE2-Diabetes extension was applied, incorporating age at diabetes diagnosis, HbA1c, and eGFR as additional predictors while retaining the same outcome definition [10]. In participants aged 70 years and older, SCORE2-OP was used [11]. For further analyses, all SCORE2-based estimates were combined and referred to collectively as SCORE2. The Framingham Risk Score (FRS10) estimates 10-year risk of coronary heart disease, cerebrovascular events, peripheral artery disease, and heart failure [12]. It was recalibrated to our study population and applied to participants aged ≥30 years. To capture longer-term risk in younger adults, the 30-year Framingham Risk Score (FRS30) was used, estimating risk of hard coronary heart disease (coronary death, myocardial infarction) and fatal or non-fatal stroke in individuals aged 20–59 years [13].

Anthropometric, clinical, and lifestyle variables were assessed using standardised protocols. Sex was self-reported with options female or male. Post menopause was defined for women as absence of menses for more than 6 months after age 45 or age above 60 years. Body mass index (BMI) was calculated as weight divided by height squared (kg/m²) and categorised as underweight (<18.5), normal weight (<25), overweight (<30), or obese (≥30 kg/m²). Blood pressure was measured twice after 5 minutes of rest in a seated position and the second reading was used. Hypertension was defined as systolic blood pressure ≥140 mmHg, diastolic blood pressure ≥90 mmHg, or self-reported physician diagnosis. Blood samples were analysed using standardised laboratory methods [14] . Measurements included haemoglobin A1c (HbA1c), high-sensitivity C-reactive protein (hsCRP), creatinine, cystatin C, total cholesterol, high-density lipoprotein (HDL), low-density lipoprotein (LDL), triglycerides, urea, uric acid, alanine aminotransferase (ALT), aspartate aminotransferase (AST), gamma-glutamyltransferase (GGT), Alkaline phosphatase (ALP). All laboratory markers were sex-specifically winsorized to the 99%-percentile before the analysis. Estimated glomerular filtration rate (eGFR) was calculated using the 2021 CKD-EPI equation based on creatinine and cystatin C [15]. Diabetes was defined by self-report or HbA1c ≥6.5% according to WHO criteria [16], and hypercholesterolaemia as LDL ≥130 mg/dL or self-reported diagnosis. History of cardiovascular disease was defined by self-reported diagnoses, including myocardial infarction, coronary artery disease, angina pectoris, heart failure, arrhythmia, peripheral artery disease, or stroke. Medication use (antihypertensive, antidiabetic, or lipid-lowering) was self-reported. Lifestyle factors included physical activity assessed by the Global Physical Activity Questionnaire, with participants classified physically active or inactive according to WHO recommendations [16], alcohol intake expressed as grams per day, and smoking status (never, former, current).

*Outcome and Covariate Assessment in UKB*

In UKB, CVD risk scores were not calculated due to the substantial time between baseline laboratory measurements and imaging visit. Instead, prospective analyses were based on incident clinical outcomes. Outcomes comprising diabetes, myocardial infarction, ischemic stroke, MACE, and hypertension were ascertained using UKB data fields 41270 and 41271, which include ICD-9 and ICD-10 diagnoses from linked hospital inpatient records. Incident diabetes was defined using ICD-10 codes E10–E14 or ICD-9 code 250. Major adverse cardiovascular events (MACE) comprised myocardial infarction (ICD-10 I21-22; ICD-9 410-411) or ischemic stroke (ICD-10 I63; ICD-9 433-434) or death from major cardiovascular diseases (ICD-10 I1-6 and I00-I78). Mortality from major cardiovascular diseases was derived through linkage to national death registries (data field 40001). Incident hypertension was defined as presence of ICD-10 codes I10 to I15 or ICD-9 codes 401 to 405. Follow-up time was calculated from the date of imaging visit to the first occurrence of an outcome of interest, death, loss to follow-up, 31 October 2022 (censoring date for ICD-based morbidity outcomes), or May 25, 2023 (download date of mortality data), whichever came first.

Participants´ sex was obtained from National Health Service (NHS) registry records at recruitment and, where applicable, updated by self-report at baseline with options female or male. At the MRI visit, participants completed a standardized questionnaire and underwent anthropometric assessments. Weight, height, and waist circumference were measured at the imaging visit. BMI was calculated as stated before for NAKO participants. Smoking status and alcohol consumption were self-reported. Smoking was dichotomized into “ever smokers” (former or current) and “never smokers.” Regular alcohol consumption was defined as alcohol intake reported as “daily or almost daily,” “three or four times a week,” or “once or twice a week.” Prevalent diabetes and history of cancer were self-reported at the imaging visit. History of stroke (ICD-10 I63; ICD-9 433-434) and myocardial infarction (ICD-10 I21-22; ICD-9 410-411) was ascertained based on recorded event dates from electronic health records prior to the imaging visit or self-report. History of hypertension was defined as ICD-10 codes I10-15 or ICD-9 codes 401-405 in electronic health records prior to the imaging visit.

### Supplementary Tables

**Supplementary Table 1:** Number of missings in adipose tissue variables.

| | Number of missings (Percentage) | |
|---|---|---|
| | NAKO<br>N = 30,868 | UK Biobank<br>N = 36,297 |
| BMFF L1 | 741 (2.40%) | 0 (0%) |
| BMFF L2 | 741 (2.40%) | 0 (0%) |
| VAT | 819 (2.65%) | 1 (0%) |
| SAT | 830 (2.68%) | 0 (0%) |
| CFF | 590 (1.91%) | 13 (0.04%) |
| RFF | 548 (1.78%) | 104 (0.29%) |
| HFF | 1076 (3.48%) | 11 (0.03%) |
| SMFF | 930 (3.01%) | 3 (0.01%) |
| IMAT | 931 (3.01%) | 0 (0%) |
| PFF | 573 (1.86%) | 69 (0.19%) |

Abbreviations. BMFF L1/L2: bone marrow fat fraction at vertebrae L1/L2, VAT: visceral adipose tissue, SAT: subcutaneous adipose tissue, CFF: Cardiac fat fraction, RFF: renal sinus fat fraction, HFF: hepatic fat fraction, SMFF: skeletal muscle fat fraction, IMAT: intramuscular adipose tissue, PFF: pancreatic fat fraction

**Supplementary Table 2:** Characteristics of NAKO and UK Biobank study populations stratified by sex.

| | NAKO | | | | UK Biobank | | |
|---|---|---|---|---|---|---|---|
| | All (N=29,314) | Men (N=16,325) | Women (N=12,989) | | All (N=36,109) | Men (N=17,453) | Women (N=18,656) |
| **Demographics** | | | | | | | |
| Age, years | 48.2 (12.3) | 47.9 (12.4) | 48.7 (12.2) | Age, years | 65.0 (7.8) | 65.7 (7.9) | 64.4 (7.6) |
| Post menopause | | | 5515 (47.3%) | | | | |
| **Anthropometry** | | | | | | | |
| BMI, kg/m² | 26.5 (4.7) | 27.0 (4.1) | 26.0 (5.2) | BMI, kg/m² | 26.2 (4.3) | 26.8 (3.9) | 25.7 (4.7) |
| Underweight | 235 (0.8%) | 65 (0.4%) | 170 (1.3%) | Underweight | 390 (1.1%) | 47 (0.3%) | 343 (1.8%) |
| Normal weight | 11,917 (40.6%) | 5,469 (33.4%) | 6,448 (49.7%) | Normal weight | 16,480 (45.8%) | 6777 (39.0%) | 9703 (52.2%) |
| Overweight | 11,312 (38.6%) | 7,486 (45.9%) | 3,826 (29.5%) | Overweight | 13,690 (38.1%) | 7878 (45.3%) | 5812 (31.3%) |
| Obese | 5,850 (20.0%) | 3,305 (20.2%) | 2,545 (19.6%) | Obese | 5,396 (15.0%) | 2671 (15.4%) | 2725 (14.7%) |
| Waist circumference, cm | 90.9 (13.6) | 95.5 (12.2) | 85.2 (13.0) | Waist circumference, cm | 88.6 (12.7) | 94.5 (10.7) | 83.0 (11.8) |
| **Lifestyle factors** | | | | | | | |
| Never Smoker | 14,389 (50.1%) | 7,509 (46.7%) | 6,894 (54.3%) | Never Smoker | 16,570 (46.6%) | 7,381 (42.9%) | 9,189 (50.1%) |
| Former Smoker | 8,897 (31.0%) | 5,406 (33.6%) | 3,508 (27.6%) | Ever Smoker | 18,992 (53.4%) | 9,830 (57.1%) | 9,162 (49.9%) |
| Current Smoker | 5,455 (19.0%) | 3,162 (19.7%) | 2,299 (18.1%) | | | | |
| Alcohol consumption, g/day | 4.6 [1.3, 13.3] | 7.3 [2.3, 17.4] | 2.7 [0.8, 8.7] | Regular alcohol consumption | 25,431 (71.3%) | 13,434 (77.9%) | 11,997 (65.1%) |
| Physically active | 23,516 (85.8%) | 13,161 (86.3%) | 10,377 (85.1%) | | | | |
| **CVD risk scores** | | | | | | | |
| SCORE2, % | 3.6 [2.0, 6.3] | 4.8 [3.0, 7.8] | 2.2 [1.2, 4.2] | | | | |
| FRS 10, % | 9.5 [4.7, 18.0] | 13.7 [7.6, 23.7] | 5.8 [3.2, 10.8] | | | | |
| FRS 30, % | 22.5 [11.5, 36.7] | 28.8 [16.1, 42.9] | 16.1 [8.3, 26.2] | | | | |
| **History of disease** | | | | | | | |
| Diabetes | 1,453 (5.0%) | 858 (5.3%) | 595 (4.6%) | Diabetes | 1,136 (3.2%) | 731 (4.2%) | 405 (2.2%) |
| Hypertension | 11,103 (37.9%) | 7,035 (43.0%) | 4,098 (31.6%) | Hypertension | 5,410 (15.0%) | 3,225 (18.6%) | 2,185 (11.8%) |
| Hypercholesterolemia | 7,080 (24.2%) | 4096 (25.1%) | 2984 (23.0%) | | | | |
| CVD | 2,875 (9.8%) | 1,522 (9.3%) | 1,353 (10.4%) | | | | |
| Myocardial infarction | 94 (0.3%) | 71 (0.4%) | 23 (0.2%) | Myocardial infarction | 558 (1.6%) | 462 (2.7%) | 96 (0.5%) |
| Stroke | 276 (0.9%) | 163 (1.0%) | 113 (0.9%) | Stroke | 166 (0.5%) | 120 (0.7%) | 46 (0.2%) |

| | | | | | | | |
|---|---|---|---|---|---|---|---|
| | | | | MACE | 1,339 (3.7%) | 1,018 (5.9%) | 321 (1.7%) |
| Cancer | 1,541 (5.3%) | 676 (4.2%) | 865 (6.7%) | Cancer | 4,293 (12.1%) | 1,974 (11.5%) | 2,319 (12.6%) |
| **Incident disease** | | | | | | | |
| | | | | Diabetes | 584 (1.7%) | 386 (2.3%) | 198 (1.1%) |
| | | | | Hypertension | 2667 (8.7%) | 1,536 (10.9%) | 1,131 (6.9%) |
| | | | | Myocardial infarction | 307 (0.9%) | 221 (1.3%) | 86 (0.5%) |
| | | | | Stroke | 226 (0.6%) | 136 (0.8%) | 90 (0.5%) |
| | | | | MACE | 613 (1.7%) | 419 (2.4%) | 194 (1.0%) |
| | | | | All-cause mortality | 625 (1.7%) | 406 (2.3%) | 219 (1.2%) |
| **Follow-up time** | | | | | | | |
| | | | | Diabetes, years | 4.2 [3.3, 5.5] | 4.2 [3.3, 5.5] | 4.2 [3.4, 5.6] |
| | | | | Hypertension, years | 4.1 [3.2, 5.4] | 4.1 [3.2, 5.4] | 4.1 [3.3, 5.4] |
| | | | | Myocardial infarction, years | 4.2 [3.4, 5.6] | 4.2 [3.3, 5.5] | 4.2 [3.4, 5.6] |
| | | | | Stroke, years | 4.2 [3.4, 5.6] | 4.2 [3.3, 5.5] | 4.2 [3.4, 5.6] |
| | | | | MACE, years | 4.2 [3.4, 5.6] | 4.2 [3.3, 5.5] | 4.2 [3.4, 5.6] |
| | | | | All-cause mortality, years | 4.8 [3.9, 6.1] | 4.8 [3.9, 6.1] | 4.8 [3.9, 6.1] |
| **Adipose tissue** | | | | | | | |
| BMFF L1, % | 48.4 (11.5) | 48.7 (10.6) | 48.0 (12.6) | BMFF L1, % | 55.3 (9.4) | 53.5 (9.2) | 56.9 (9.3) |
| BMFF L2, % | 54.8 (14.6) | 56.3 (13.2) | 53.0 (15.9) | BMFF L2, % | 56.2 (9.4) | 54.7 (9.2) | 57.7 (9.4) |
| VAT, l | 3.0 [1.7, 4.9] | 4.2 [2.6, 5.9] | 1.9 [1.2, 3.2] | VAT, l | 3.2 [1.8, 4.9] | 4.6 [3.1, 6.2] | 2.2 [1.3, 3.3] |
| SAT, l | 12.0 [8.9, 16.1] | 10.9 [8.2, 14.2] | 13.7 [10.1, 18.5] | SAT, l | 12.8 [9.9, 16.7] | 11.4 [9.0, 14.4] | 14.6 [11.2, 18.7] |
| CFF, % | 23.2 (5.5) | 23.3 (5.6) | 23.0 (5.3) | CFF, % | 19.3 (3.7) | 19.5 (3.6) | 19.2 (3.7) |
| RFF, % | 49.7 (11.7) | 53.2 (10.9) | 45.4 (11.2) | RFF, % | 48.7 (10.6) | 53.1 (9.3) | 44.6 (10.1) |
| HFF, % | 3.5 [2.4, 6.6] | 4.2 [2.9, 8.3] | 2.7 [2.0, 4.7] | HFF, % | 4.4 [3.9, 5.7] | 4.7 [4.1, 6.4] | 4.1 [3.7, 4.9] |
| SMFF, % | 17.3 (4.0) | 16.3 (3.8) | 18.5 (4.0) | SMFF, % | 18.4 (3.7) | 17.1 (3.4) | 19.6 (3.5) |
| IMAT, $dm^3$ | | | | IMAT, $dm^3$ | 20.4 (6.9) | 20.7 (6.2) | 20.0 (7.6) |
| PFF, % | 7.9 [5.6, 12.3] | 9.2 [6.6, 14.4] | 6.5 [4.8, 9.7] | PFF, % | 10.0 [7.6, 13.9] | 11.6 [8.9, 16.1] | 8.7 [6.7, 11.7] |
| **Clinical marker** | | | | | | | |
| Systolic Blood Pressure, mmHg | 127.5 (15.9) | 131.0 (14.6) | 123.1 (16.3) | | | | |
| Diastolic Blood Pressure, mmHg | 78.9 (9.9) | 80.4 (9.8) | 77.0 (9.7) | | | | |
| HbA1c, mmol/mol | 35.8 (5.3) | 36.0 (5.6) | 35.7 (4.8) | | | | |
| Total cholesterol, mg/dl | 206.1 (39.9) | 204.3 (39.6) | 208.5 (40.2) | | | | |

| | | | |
|---|---|---|---|
| HDL, mg/dl | 59.0 (15.9) | 52.6 (12.6) | 67.1 (15.9) |
| LDL, mg/dl | 128.4 (33.9) | 130.6 (33.3) | 125.6 (34.3) |
| Triglycerides, mg/dl | 116.9 [82.4, 173.6] | 132.9 [92.1, 197.5] | 101.9 [74.4, 143.5] |
| hsCRP, mg/l | 1.0 [0.5, 2.1] | 0.9 [0.5, 1.8] | 1.1 [0.5, 2.7] |
| eGFR, ml/min/1.73m² | 102.7 (15.0) | 103.5 (14.7) | 101.7 (15.1) |
| Creatinine, µmol/l | 73.5 (13.6) | 80.6 (11.9) | 64.6 (9.7) |
| Cystatin C, mg/l | 0.8 (0.1) | 0.8 (0.1) | 0.8 (0.1) |
| Urea, mmol/l | 5.1 (1.3) | 5.4 (1.3) | 4.8 (1.3) |
| Uric acid, µmol/l | 294.9 (76.9) | 334.3 (65.8) | 245.1 (58.9) |
| ALP, U/l | 70.2 (18.9) | 71.5 (17.8) | 68.5 (20.1) |
| GGT, U/l | 26.9 [19.8, 38.3] | 31.7 [24.0, 45.5] | 22.2 [16.8, 28.7] |
| AST, U/l | 22.2 [18.0, 26.9] | 24.0 [19.8, 28.7] | 19.8 [16.8, 24.0] |
| ALT, U/l | 26.9 [20.4, 35.9] | 31.1 [24.6, 41.3] | 22.2 [18.0, 28.1] |
| **Medication** | | | |
| Antihypertensive | 5,740 (19.9%) | 3,344 (20.8%) | 2,396 (18.7%) |
| Antidiabetic | 858 (3.0%) | 563 (3.5%) | 299 (2.3%) |
| Lipid-lowering | 1,937 (6.7%) | 1244 (7.7%) | 693 (5.4%) |

Values are given as mean (standard deviation) or median [interquartile range] for continuous values and counts (percentage) for categorical values. Abbreviations. BMI: Body mass index, CVD: cardiovascular disease, MACE: major adverse cardiovascular events, BMFF L1/L2: bone marrow fat fraction at vertebrae L1/L2, VAT: visceral adipose tissue, SAT: subcutaneous adipose tissue, CFF: Cardiac fat fraction, RFF: renal sinus fat fraction, HFF: hepatic fat fraction, SMFF: skeletal muscle fat fraction, IMAT: intramuscular adipose tissue, PFF: pancreatic fat fraction, HbA1c: haemoglobin A1c, HDL: high-density lipoprotein, LDL: low-density lipoprotein, hsCRP: high-sensitivity C-reactive protein, eGFR: estimated glomerular filtration rate, ALP: Alkaline phosphatase, GGT: gamma-glutamyltransferase, AST: aspartate aminotransferase, ALT: alanine aminotransferase.

**Supplementary Table 3:** Cluster stability assessment in NAKO.

Cluster stability was assessed by drawing 1000 bootstrap samples with replacement from the NAKO data set and repeating the clustering on each of the bootstrapped samples. The Jaccard Index, a partition similarity index that measures to which extent the clusters of the bootstrapped sample coincide with the original ones, is computed cluster-wise: For each bootstrap sample, the original cluster was compared with the most similar cluster in the resampled solution, and the Jaccard coefficient was calculated as the proportion of shared observations among all observations belonging to either cluster. The mean of Jaccard Indices for each cluster is used as an index of stability. Valid, stable cluster should yield a mean Jaccard similarity value of 0.75 or more and values of 0.85 or above indicate highly stable clusters. A value < 0.5 indicates highest instability, referred to as dissolved cluster [17]. Mean Jaccard Index and percentages of dissolved clusters are presented.

| Cluster | Mean Jaccard Index | Percentage of dissolved cluster |
|---|---|---|
| I | 0.95 | 0% |
| II | 0.87 | 1.4% |
| III | 0.81 | 2.3% |
| IV | 0.59 | 47% |
| V | 0.77 | 2.2% |

**Supplementary Table 4:** Characteristics of NAKO and UK Biobank populations stratified by subphenotype.

| | Subphenotypes in NAKO | | | | | | Subphenotypes in UK Biobank | | | | |
|---|---|---|---|---|---|---|---|---|---|---|---|
| | I<br>N = 6,869 | II<br>N = 9,825 | III<br>N = 7,514 | IV<br>N = 2,453 | V<br>N = 2,653 | | I<br>N = 3,837 | II<br>N = 13,904 | III<br>N = 12,734 | IV<br>N = 1,891 | V<br>N = 3,743 |
| **Demographics** | | | | | | | | | | | |
| Women | 3,227 (47.0%) | 4,373 (44.5%) | 3,249 (43.2%) | 915 (37.3%) | 1,225 (46.2%) | Women | 1,590 (41.4%) | 7,358 (52.9%) | 6,805 (53.4%) | 994 (52.6%) | 1,909 (51.0%) |
| Age, years | 36.5 (10.5) | 47.3 (10.0) | 55.2 (8.9) | 51.4 (10.2) | 59.4 (7.9) | Age, years | 60.1 (7.6) | 63.7 (7.6) | 67.1 (7.2) | 62.2 (6.9) | 69.0 (6.7) |
| **Anthropometry** | | | | | | | | | | | |
| BMI, kg/m² | 23.2 (2.9) | 25.1 (3.3) | 27.5 (3.5) | 32.3 (4.8) | 32.3 (4.7) | BMI, kg/m² | 22.7 (2.7) | 24.1 (3.1) | 26.5 (3.4) | 31.6 (4.9) | 30.8 (4.4) |
| Underweight | 162 (2.9%) | 65 (0.7%) | 7 (0.1%) | 1 (0.0%) | 0 (0.0%) | Underweight | 162 (4.2%) | 201 (1.5%) | 26 (0.2%) | 1 (0.1%) | 0 (0%) |
| Normal weight | 5,040 (73.4%) | 5,010 (51.0%) | 1,703 (22.7%) | 81 (3.3%) | 83 (3.1%) | Normal weight | 2,999 (78.4%) | 8,823 (63.8%) | 4,299 (33.9%) | 106 (5.6%) | 253 (6.8%) |
| Overweight | 1,517 (22.1%) | 4,001 (40.7%) | 4,192 (55.8%) | 792 (32.3%) | 810 (30.5%) | Overweight | 616 (16.1%) | 4,324 (31.2%) | 6,555 (51.7%) | 678 (35.9%) | 1,517 (40.7%) |
| Obese | 150 (2.2%) | 749 (7.6%) | 1,612 (21.5%) | 1,579 (64.4%) | 1,760 (66.3%) | Obese | 46 (1.2%) | 489 (3.5%) | 1,799 (14.2%) | 1,102 (58.4%) | 1,960 (52.5%) |
| Waist circumference, cm | 79.6 (8.5) | 86.7 (9.7) | 95.3 (9.8) | 106.4 (11.3) | 108.2 (11.2) | Waist circumference, cm | 78.9 (9.1) | 83.1 (10.2) | 91.0 (10.0) | 102.6 (11.9) | 103.2 (11.0) |
| **Lifestyle factors** | | | | | | | | | | | |
| Never Smoker | 4,175 (61.4%) | 4,846 (50.2%) | 3,194 (43.6%) | 1,105 (46.1%) | 1,069 (41.6%) | Never Smoker | 2,008 (53.0%) | 6,720 (49.0%) | 5,516 (44.0%) | 860 (45.8%) | 1,466 (40.1%) |
| Former Smoker | 1,455 (21.4%) | 2,824 (29.2%) | 2,655 (36.3%) | 846 (35.3%) | 1,117 (43.5%) | Ever Smoker | 1,782 (47.0%) | 6,989 (51.0%) | 7,010 (56.0%) | 1,017 (54.2%) | 2,194 (59.9%) |
| Current Smoker | 1,166 (17.2%) | 1,991 (20.6%) | 1,469 (20.1%) | 447 (18.6%) | 382 (14.9%) | | | | | | |
| Alcohol consumption, g/day | 4.1 [1.3, 10.1] | 4.7 [1.4, 12.7] | 5.5 [1.4, 16.2] | 4.1 [1.0, 14.8] | 4.8 [1.1, 16.3] | Regular alcohol consumption | 2,700 (71.2%) | 10,010 (72.8%) | 9,073 (72.2%) | 1,153 (61.3%) | 2,495 (67.8%) |
| Physically active | 5,929 (91.1%) | 7,848 (85.6%) | 5,870 (84.0%) | 1,857 (80.2%) | 2,012 (82.4%) | | | | | | |
| **History of disease** | | | | | | | | | | | |
| Diabetes | 108 (1.6%) | 192 (2.0%) | 355 (4.7%) | 389 (15.9%) | 409 (15.5%) | Diabetes | 32 (0.8%) | 176 (1.3%) | 435 (3.4%) | 146 (7.7%) | 347 (9.3%) |
| Hypertension | 967 (14.1%) | 2,834 (28.9%) | 3,796 (50.6%) | 1,619 (66.2%) | 1,887 (71.3%) | Hypertension | 212 (5.5%) | 1,297 (9.4%) | 2,287 (18.0%) | 410 (21.7%) | 1,204 (32.3%) |
| Hypercholesterolemia | 547 (8.0%) | 1,979 (20.1%) | 2,546 (33.9%) | 922 (37.6%) | 1,086 (41.0%) | | | | | | |
| CVD | 362 (5.3%) | 784 (8.0%) | 943 (12.5%) | 290 (11.8%) | 496 (18.7%) | | | | | | |
| Myocardial infarction | 3 (0.0%) | 17 (0.2%) | 41 (0.5%) | 12 (0.5%) | 21 (0.8%) | Myocardial infarction | 35 (0.9%) | 130 (0.9%) | 265 (2.1%) | 21 (1.1%) | 107 (2.9%) |
| Stroke | 14 (0.2%) | 51 (0.5%) | 108 (1.4%) | 38 (1.6%) | 65 (2.5%) | Stroke | 8 (0.2%) | 46 (0.3%) | 63 (0.5%) | 9 (0.5%) | 40 (1.1%) |
| | | | | | | MACE | 76 (2.0%) | 326 (2.4%) | 588 (4.6%) | 63 (3.3%) | 286 (7.7%) |
| Cancer | 166 (2.4%) | 413 (4.2%) | 566 (7.6%) | 167 (6.8%) | 229 (8.7%) | Cancer | 345 (9.1%) | 1,511 (11.0%) | 1,617 (12.9%) | 214 (11.4%) | 606 (16.5%) |
| **Clinical marker** | | | | | | | | | | | |
| Systolic Blood Pressure, mmHg | 120.8 (13.1) | 125.6 (14.9) | 131.5 (16.4) | 133.4 (15.1) | 134.3 (17.0) | | | | | | |
| Diastolic Blood Pressure, mmHg | 73.9 (8.6) | 78.4 (9.4) | 81.2 (9.6) | 84.2 (9.4) | 82.2 (10.0) | | | | | | |
| HbA1c, mmol/mol | 33.3 (3.8) | 34.9 (4.0) | 36.8 (4.8) | 39.5 (7.3) | 39.6 (6.7) | | | | | | |
| Total cholesterol, mg/dl | 183.1 (34.2) | 207.1 (37.1) | 219.6 (38.8) | 211.9 (39.2) | 218.6 (41.3) | | | | | | |
| HDL, mg/dl | 62.2 (15.1) | 61.1 (16.3) | 58.4 (15.9) | 48.4 (12.5) | 54.7 (13.8) | | | | | | |
| LDL, mg/dl | 109.0 (28.8) | 128.8 (31.9) | 139.5 (33.0) | 135.6 (32.9) | 139.2 (34.3) | | | | | | |

| | | | | | |
|---|---|---|---|---|---|
| Triglycerides, mg/dl | 87.7 [65.5, 119.1] | 107.2 [77.9, 155.9] | 134.6 [98.3, 193.1] | 191.3 [132.9, 275.7] | 160.3 [117.8, 222.3] |
| hsCRP, mg/l | 0.6 [0.3, 1.1] | 0.8 [0.4, 1.6] | 1.2 [0.7, 2.4] | 2.1 [1.1, 4.1] | 2.2 [1.1, 4.1] |
| eGFR, ml/min/1.73m² | 111.0 (13.1) | 104.1 (13.3) | 98.0 (14.2) | 99.1 (15.1) | 92.8 (15.2) |
| Creatinine, µmol/l | 72.6 (13.0) | 73.4 (13.2) | 73.9 (13.9) | 75.0 (14.3) | 73.9 (14.6) |
| Cystatin C, mg/l | 0.8 (0.1) | 0.8 (0.1) | 0.8 (0.1) | 0.9 (0.1) | 0.9 (0.1) |
| Urea, mmol/l | 4.8 (1.3) | 5.1 (1.3) | 5.3 (1.3) | 5.3 (1.2) | 5.5 (1.3) |
| Uric acid, µmol/l | 269.7 (70.6) | 282.7 (72.8) | 303.5 (74.2) | 348.0 (74.8) | 332.4 (75.2) |
| ALP, U/l | 64.4 (17.4) | 68.2 (17.7) | 73.9 (18.7) | 76.8 (20.1) | 77.5 (19.9) |
| GGT, U/l | 21.6 [16.8, 27.5] | 25.1 [19.2, 34.1] | 29.9 [22.2, 43.1] | 41.3 [29.9, 59.9] | 35.3 [26.3, 52.1] |
| AST, U/l | 21.0 [17.4, 25.1] | 21.6 [18.0, 25.7] | 22.2 [18.6, 26.3] | 27.5 [22.2, 34.1] | 22.8 [19.2, 28.1] |
| ALT, U/l | 22.8 [18.0, 29.3] | 25.1 [19.8, 32.9] | 27.5 [22.2, 35.9] | 43.7 [32.3, 59.3] | 31.1 [24.0, 40.7] |
| **Medication** | | | | | |
| Antihypertensive | 219 (3.2%) | 1,086 (11.2%) | 2,103 (28.4%) | 950 (39.3%) | 1,382 (52.9%) |
| Antidiabetic | 53 (0.8%) | 76 (0.8%) | 223 (3.0%) | 234 (9.7%) | 272 (10.4%) |
| Lipid-lowering | 111 (1.6%) | 371 (3.8%) | 764 (10.3%) | 245 (10.1%) | 446 (17.1%) |

Values are given as mean (standard deviation) or median [interquartile range] for continuous values and counts (percentage) for categorical values. Abbreviations. BMI: Body mass index, CVD: cardiovascular disease, MACE: major adverse cardiovascular events, HbA1c: haemoglobin A1c, HDL: high-density lipoprotein, LDL: low-density lipoprotein, hsCRP: high-sensitivity C-reactive protein, eGFR: estimated glomerular filtration rate, ALP: Alkaline phosphatase, GGT: gamma-glutamyltransferase, AST: aspartate aminotransferase, ALT: alanine aminotransferase.

### Supplementary Figures

**Supplementary Figure 1**: Cluster size comparison.

Comparison of relative and absolute cluster sizes in NAKO and UK Biobank (UKB). For UKB, results for both standardization approaches are presented.

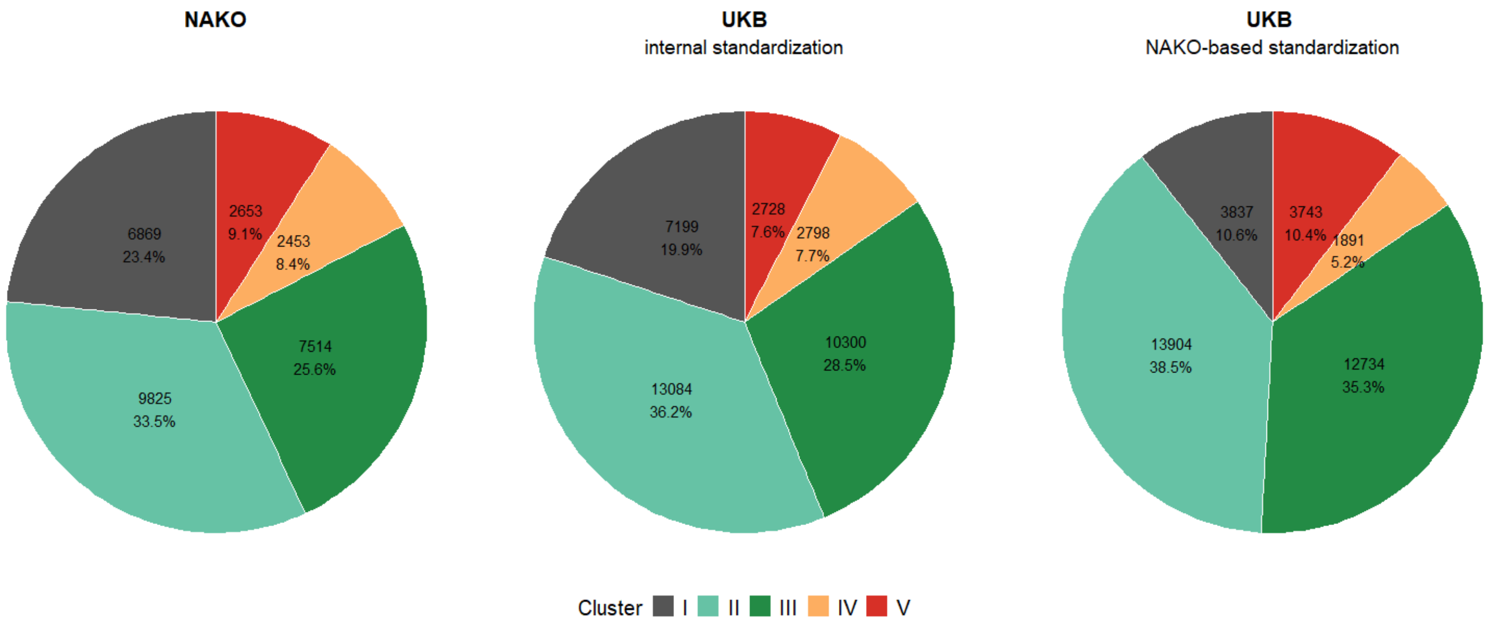

**Supplementary Figure 2**: Comparison of adipose tissue distributions across cluster and cohorts.

Cluster specific distributions of adipose tissue tissue depots in NAKO and UK Biobank (UKB). Boxplots are shown for UKB using NAKO-based standardization. Distributions were comparable when applying internal standardization and are not shown. Previously reported results from the original KORA study are included for comparison [18]. In the original study, IMAT was not available and cardiac fat measured in cm² but not in % and is therefore not presented.

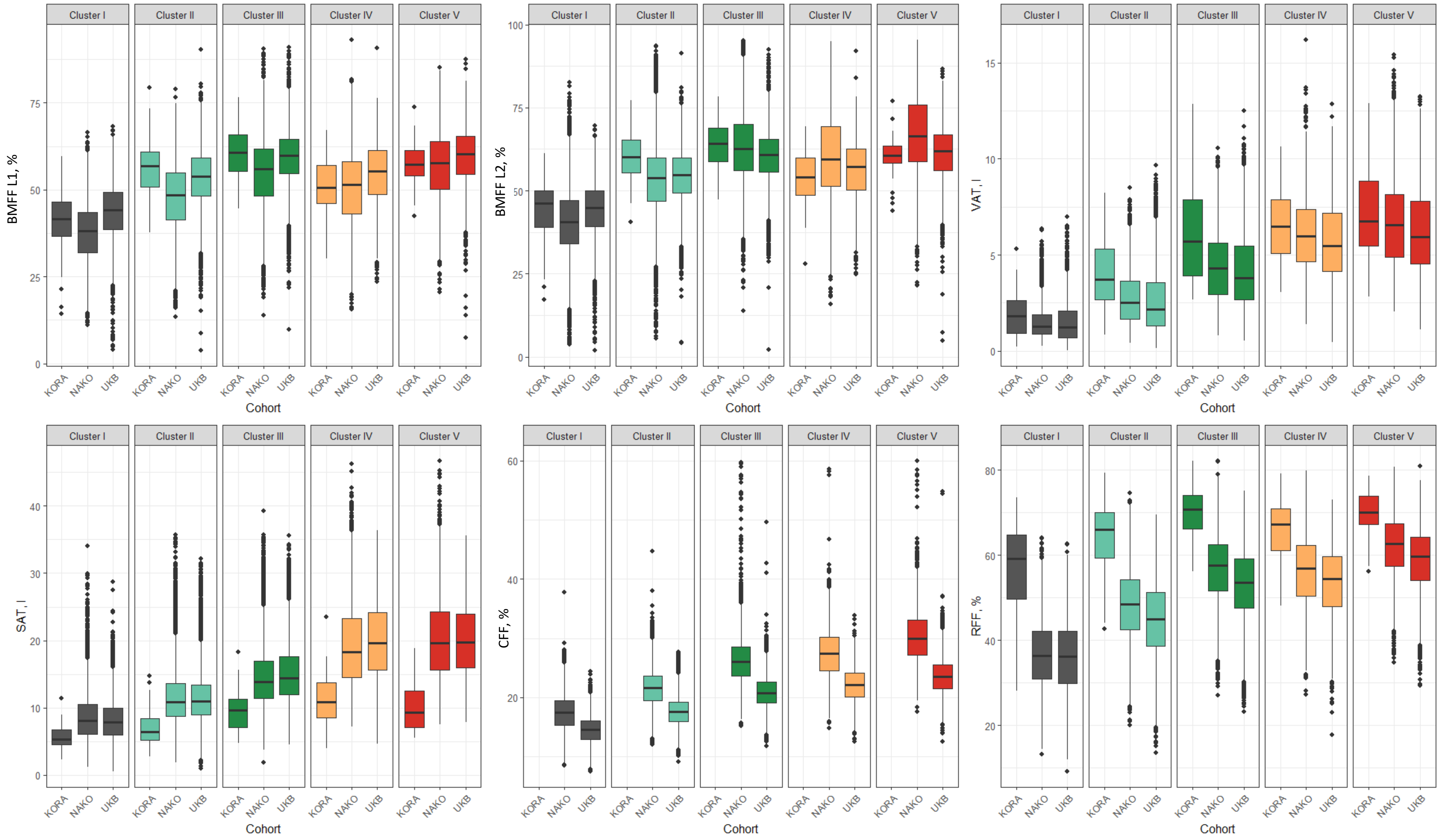

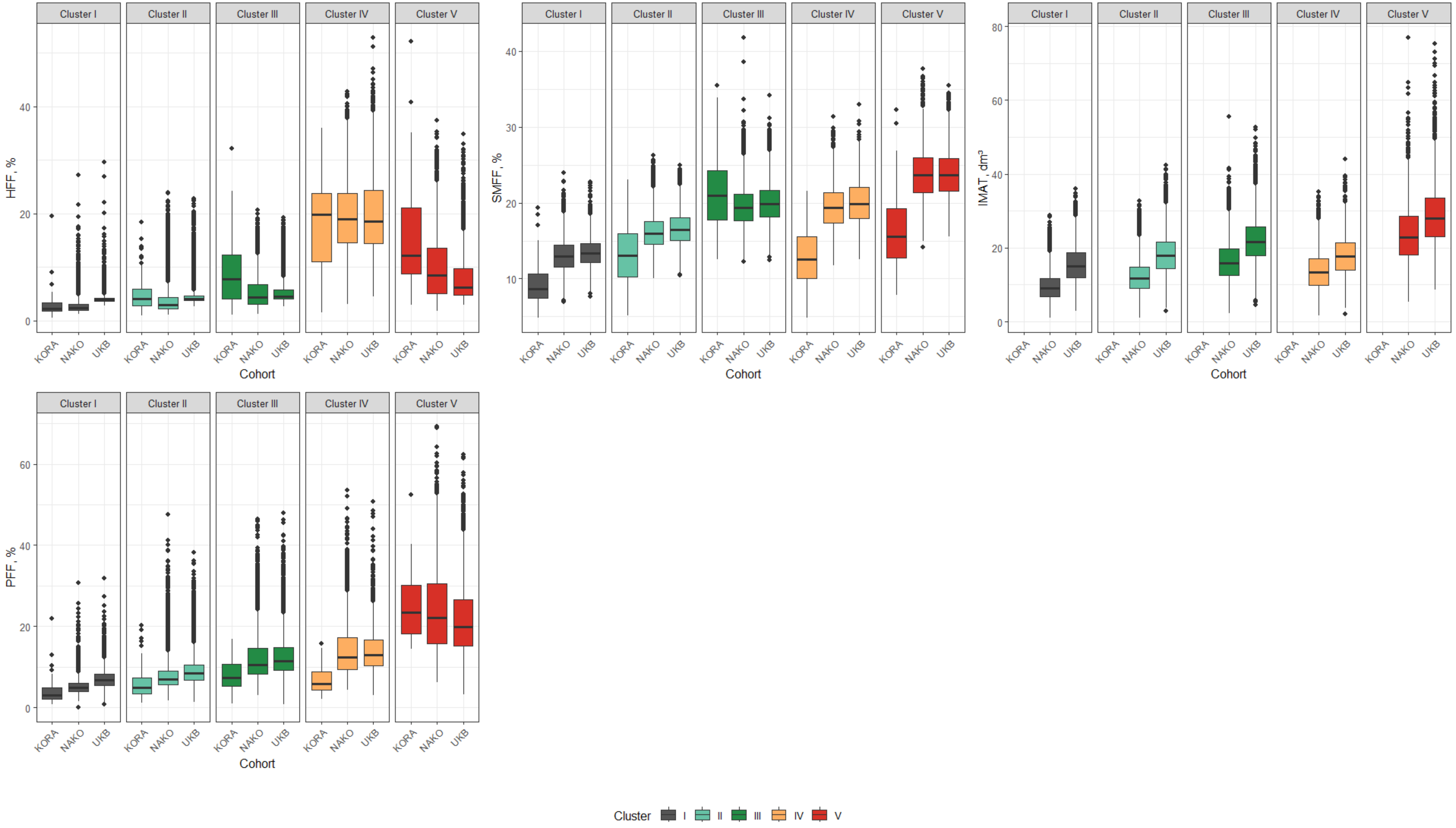

Cluster I
Cluster II
Cluster III
Cluster IV
Cluster V
HFF, %
0
20
40
SMFF, %
10
20
30
40
IMAT, dm³
0
20
40
60
80
PFF, %
0
20
40
60
KORA
NAKO
UKB
Cohort
Cluster
I
II
III
IV
V

**Supplementary Figure 3**: Principal component plots.

Separation of clusters visualized in coordinates of the two first principal components for clustering in NAKO and UK Biobank (UKB) including both standardization approaches compared to original clustering in KORA [18].

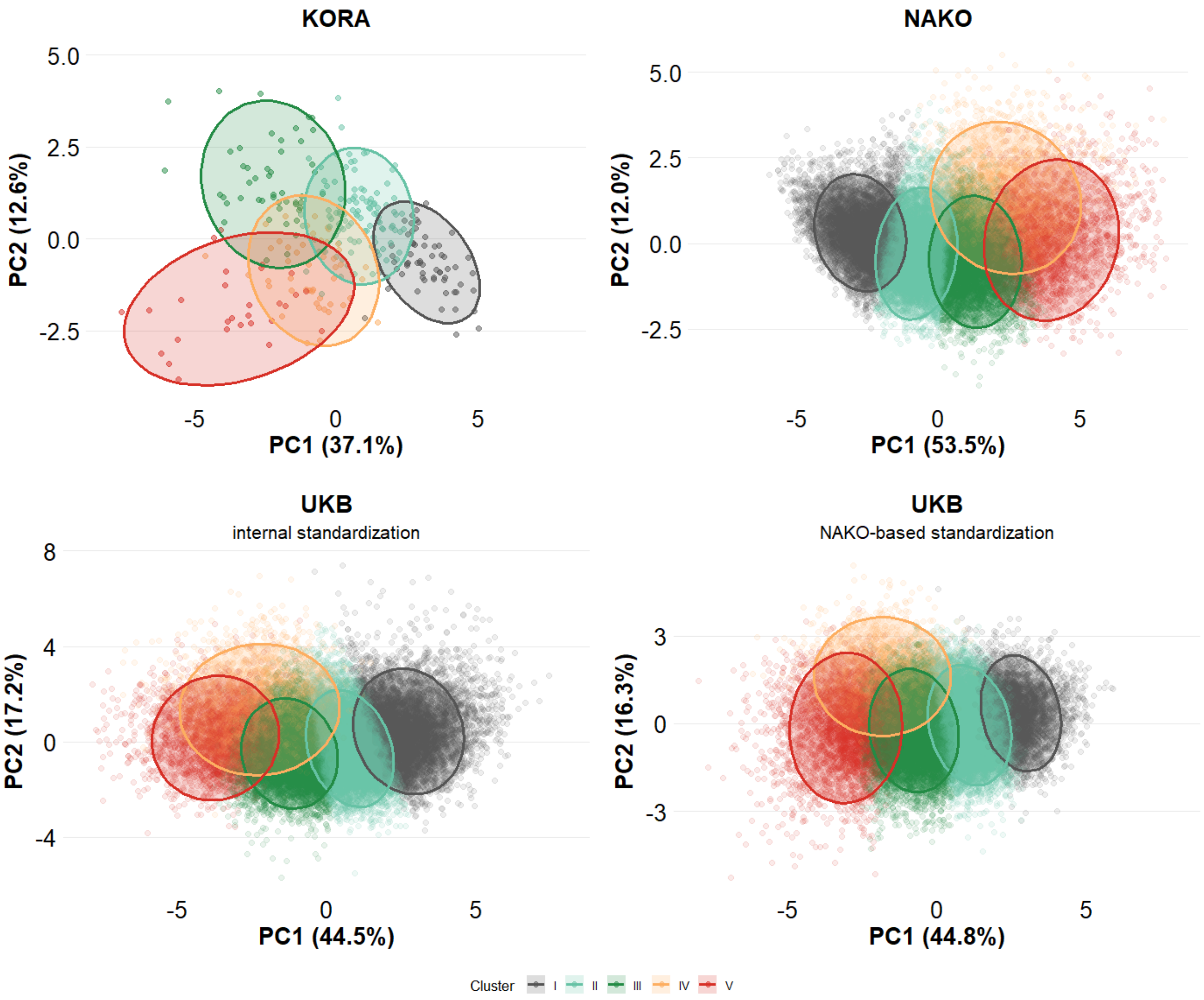

**Supplementary Figure 4**: Silhouette plots.

Silhouette plots for clustering in NAKO and UK Biobank (UKB) including both standardization approaches compared to original clustering in KORA [18]. Each bar represents the silhouette width of an individual observation, with values ranging from −1 to 1, where higher values indicate greater similarity to the assigned cluster (lower distance to individuals within one cluster) relative to other clusters (greater distance to individuals in nearest neighbouring cluster). The dashed red line denotes the average silhouette width (ASW) across all observations measuring overall cluster cohesion and separation [19].

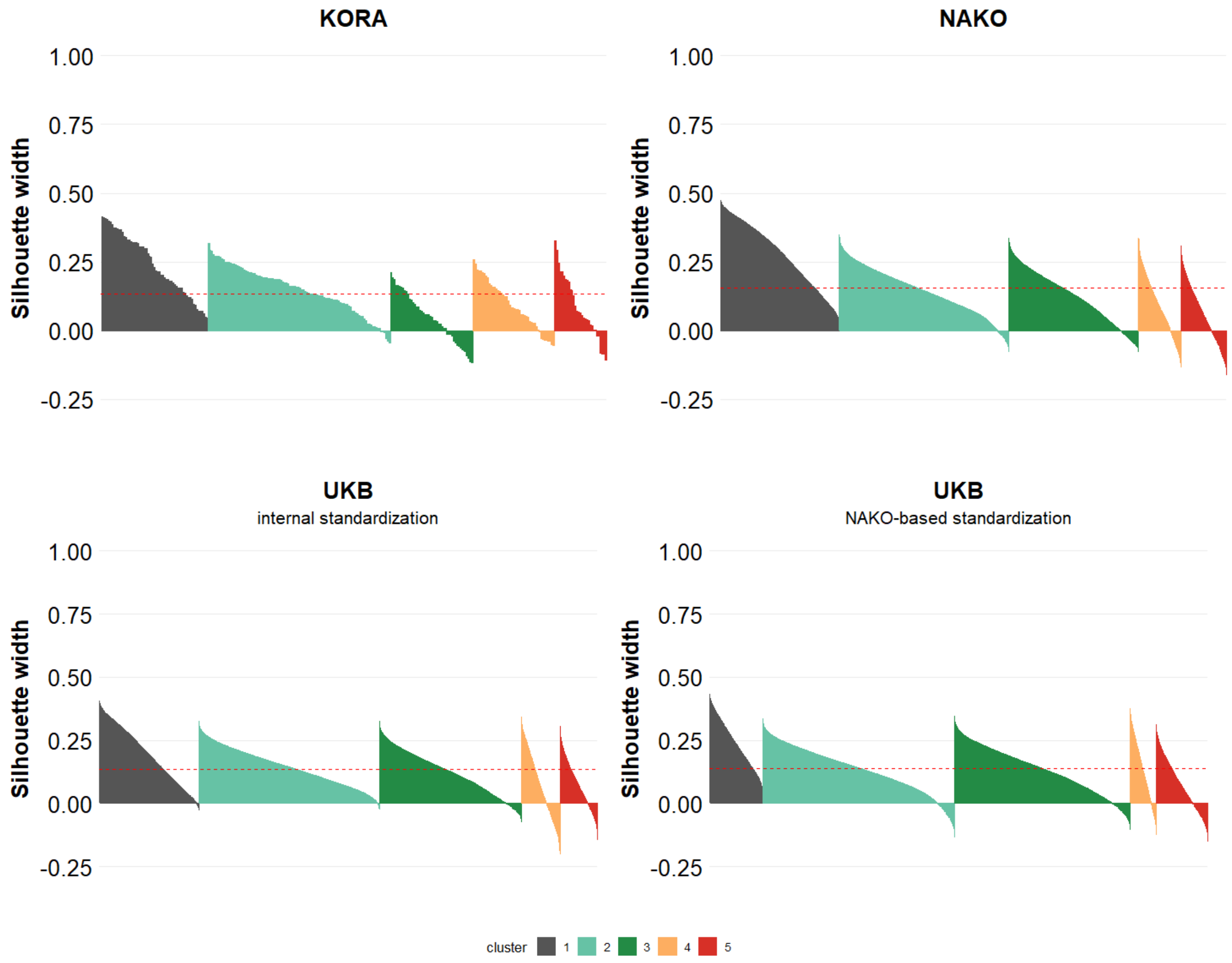

**Supplementary Figure 5:** Association between subphenotypes and CVD risk scores.

Relative cardiovascular risk increase by subphenotype for the NAKO compared to original KORA results [18]. Shown are exponentiated regression coefficients from linear regression models and CVD risk scores (SCORE2, FRS10, and FRS30) as outcomes, using subphenotype I as reference. Error bars indicate 95% confidence intervals. Models were unadjusted since risk score calculation included particularly age and sex.

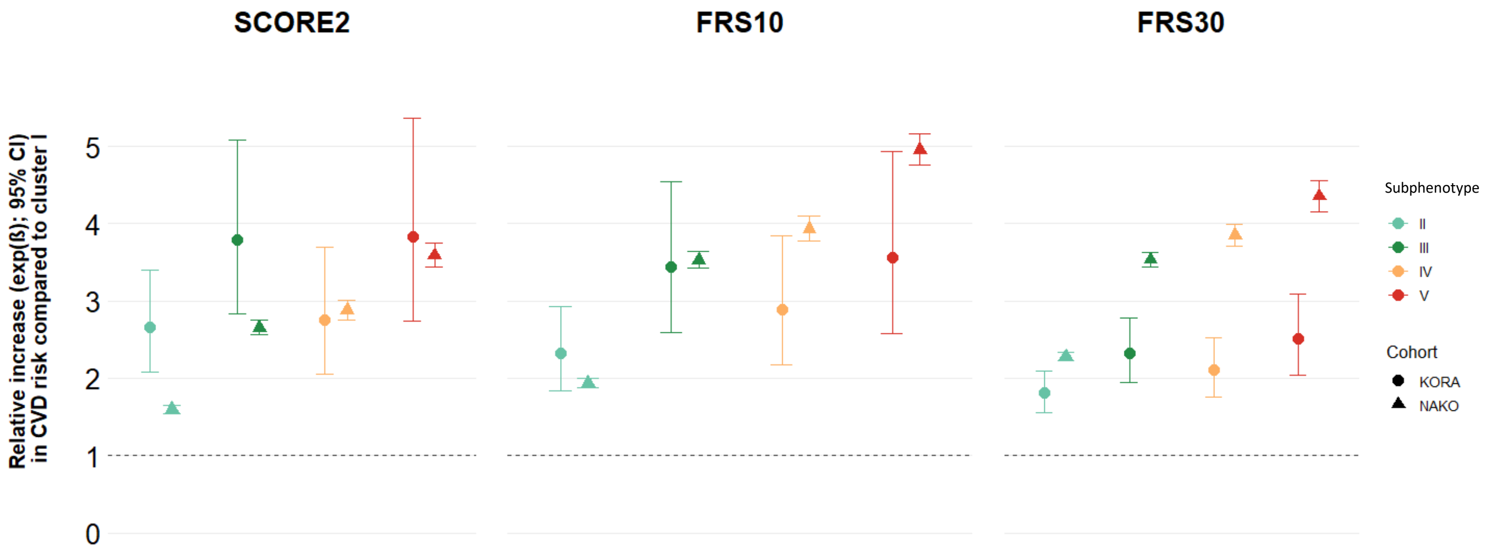

**Supplementary Figure 6**: Sex-specific radar charts in NAKO.

Sex-specific radar charts of standardized adipose tissue depots (z-scores) across the five clusters, representing body composition subphenotypes in NAKO for women based on main clustering (panel A) and clustering women separately (panel B), respectively, for men based on main clustering (panel C) and clustering men separately (panel D). The respective population means for women and men, respectively, are shown as grey polygon at 0. Colored lines represent mean values within each subphenotype. Axes range from –2 to +2, corresponding to standard deviation from the mean.

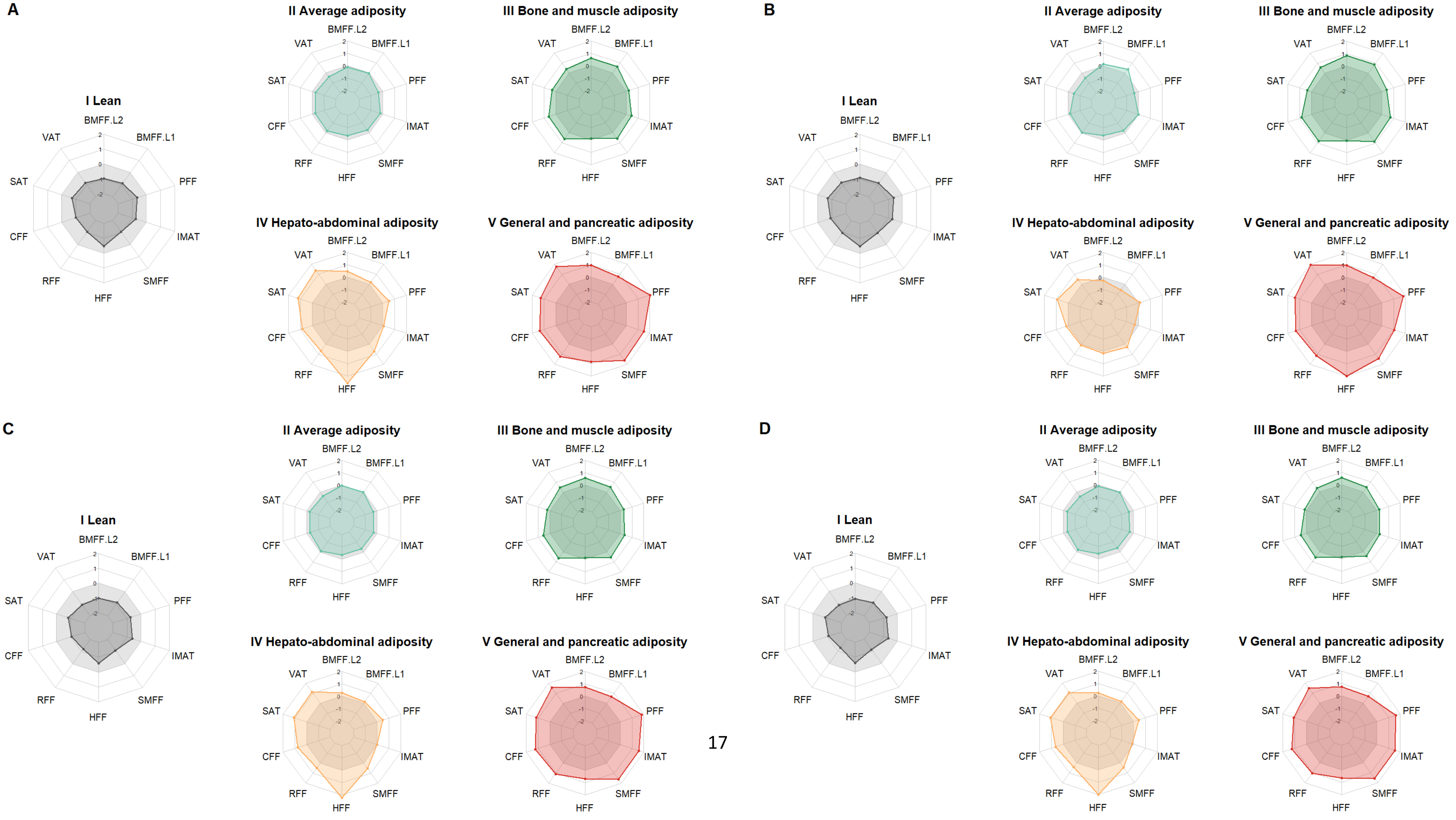

**Supplementary Figure 7**: Sex-specific forest plots in NAKO.

Relative cardiovascular risk increase by cluster for the NAKO comparing main clustering (panel A) and sex-specific clustering (panel B). Shown are exponentiated regression coefficients from sex-stratified linear regression models and CVD risk scores (SCORE2, FRS10, and FRS30) as outcomes, using cluster I as reference. Error bars indicate 95% confidence intervals. Models were unadjusted since risk score calculation included particularly age.

A

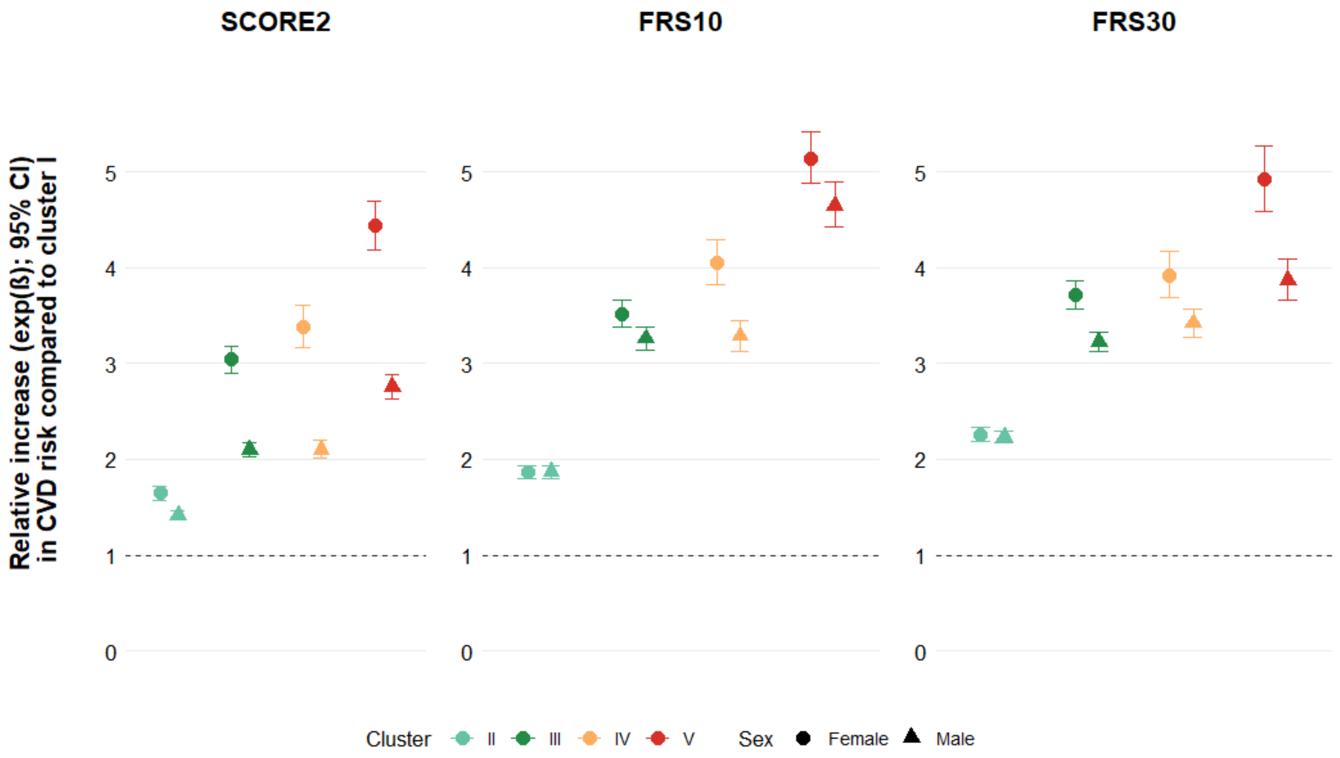

B

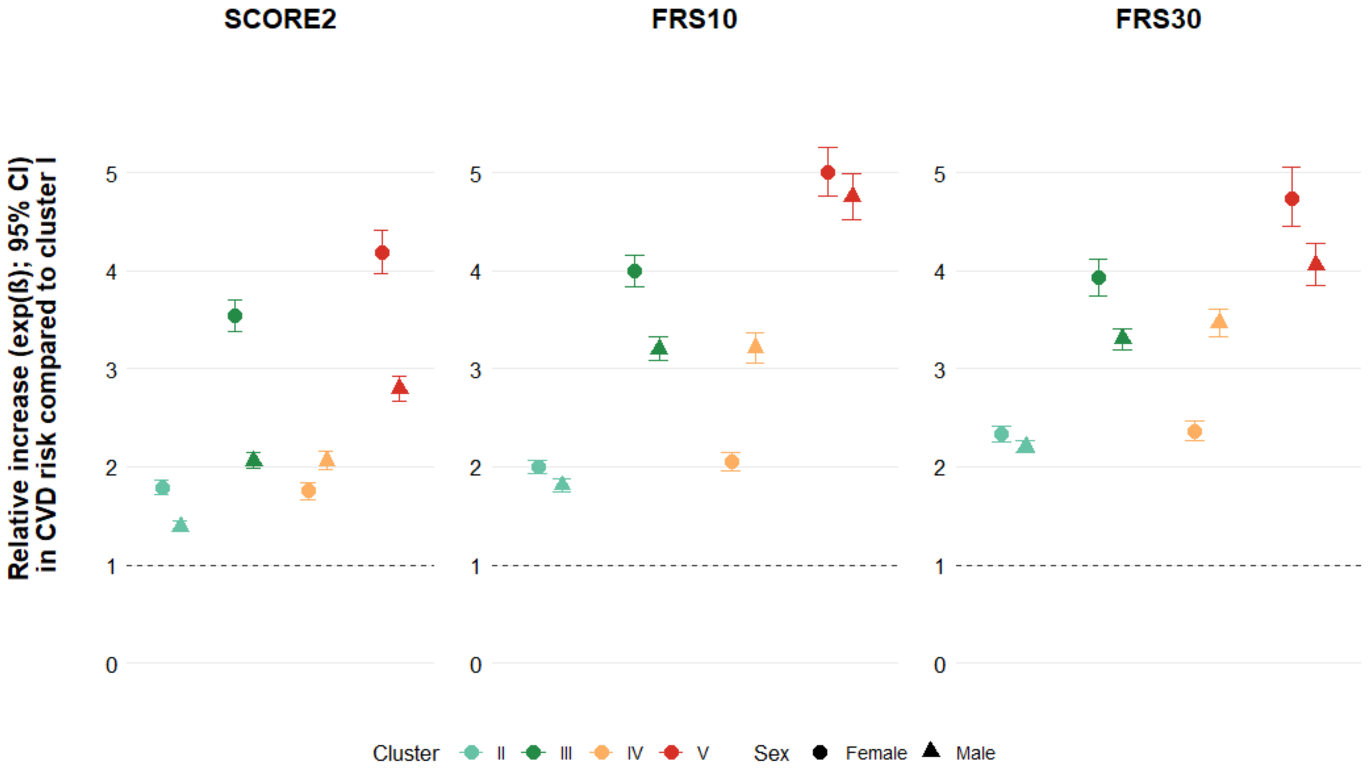

**Supplementary Figure 8:** Radar charts for women and men with increased BMI and waist circumference in NAKO.

Radar charts of standardized adipose tissue depots (z-scores) across the five clusters. Subpopulations of individuals with visceral obesity as classified by BMI 25-30 kg/m² and waist circumference for women ≥90 cm (panel A), men ≥100 cm (panel B) as defined by [20] are presented. The respective population means for women and men, respectively, are shown as grey polygon at 0. Colored lines represent mean values within each subphenotype. Axes range from –2 to +2, corresponding to standard deviation from the mean.

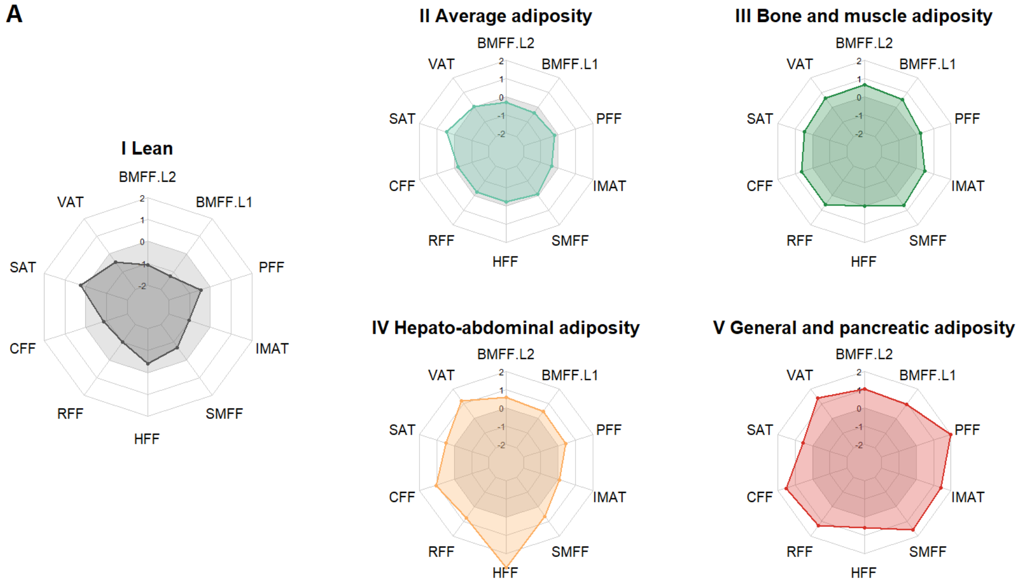

Distribution of women with BMI 25-30 kg/m² and waist circumference ≥90 cm across the subphenotypes:

| Subphenotype | I | II | III | IV | V |
|---|---|---|---|---|---|
| N | 63 | 325 | 714 | 196 | 259 |

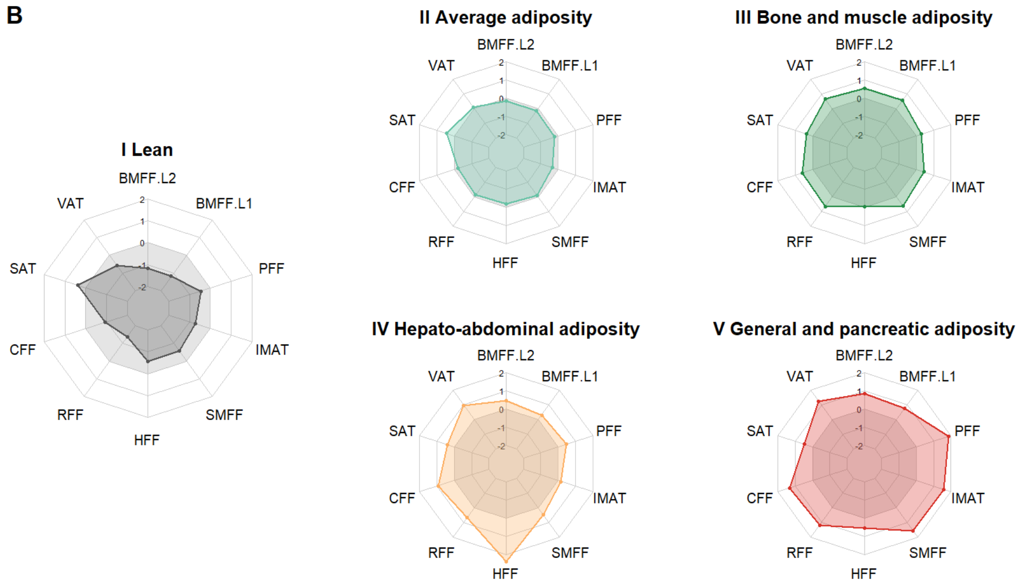

Distribution of men with BMI 25-30 kg/m² and waist circumference ≥100 cm across the subphenotypes:

| Subphenotype | I | II | III | IV | V |
|---|---|---|---|---|---|
| N | 30 | 326 | 1155 | 309 | 379 |

**Supplementary Code**

**Grune et al.**

**Body composition subphenotypes, cardiometabolic risk and incident outcomes: validation in the population-based NAKO and UK Biobank imaging cohorts**

**Validation procedure based on Ullmann et al. (https://doi.org/10.1002/widm.1444)**

**Author: Elena Grune**

**Date: 17 June 2026**

**This supplementary file provides the R code used for cluster derivation and cluster validation, and related analyses reported in the manuscript.**

**Data preparation is not included in this code. These steps were performed prior to the analyses presented here and are specific to the underlying study data, which cannot be shared because of data protection.**

**Accordingly, the code begins with a preprocessed analytical dataset and focuses on the clustering workflow, validation procedures, and generation of validation results.**

```
library(dplyr)
library(stringr)
library(ggplot2)
library(fpc)
library(ggplot2)
library(scales)
library(patchwork)
library(ggfortify)
library(gridExtra)
library(mclust)
library(ggsankey)
library(modelsummary)
library(factoextra)
library(grid)
library(cowplot)
library(scales)
library(patchwork)
library(plotly)
```

```r
library(data.table)
library(fmsb)

# dt is data frame containing NAKO data only
# dt includes particularly participant ID, age, sex, and adipose tissue (AT) variables,
# CVD risk scores, BMI, waist circumference
# First, present code for NAKO analyses

############################################################
# Define adipose tissue variables used for clustering
############################################################

AT <- c("vat_l", "sat_l", "liver_pdff", "bmat_l2", "bmat_l1",
        "smff", "imat", "pff", "ren_hil_ff", "heartfat2")

############################################################
# Sex-specific standardisation
############################################################

# females
dtf <- dt[dt$sex == 1, ]

f_scaled <- as.data.frame(scale(dtf[, AT]))
colnames(f_scaled) <- str_c(colnames(f_scaled), "_std")
dtf <- cbind(dtf, f_scaled)

# males
dtm <- dt[dt$sex == 0, ]

m_scaled <- as.data.frame(scale(dtm[, AT]))
colnames(m_scaled) <- str_c(colnames(m_scaled), "_std")
dtm <- cbind(dtm, m_scaled)

# combine
dt_fat <- rbind(dtf, dtm)

rm(f_scaled, m_scaled)

############################################################
# Main clustering
############################################################

fat <- dt_fat %>%
  select("ID_751", "age", "sex",
         "bmat_l1_std", "bmat_l2_std", "vat_l_std", "sat_l_std",
         "heartfat2_std", "ren_hil_ff_std", "liver_pdff_std",
         "smff_std", "imat_std", "pff_std")

# run cluster
fat <- fat[complete.cases(fat), ]
colSums(is.na(fat))
```

```r
set.seed(17)
k5 <- kmeans(fat[, -(1:3)], centers = 5, nstart = 25)
centroids <- k5$centers # extract cluster centroids needed for later assignment of UKB data

fat$k5 <- as.factor(k5$cluster)

# merge cluster back
final <- merge(dt_fat,
               fat[, c("ID_751", "sex", "age", "k5")],
               by = c("ID_751", "sex", "age"))

rm(fat)

############################################################
# Relabel clusters by ascending median VAT
############################################################

median_vat <- tapply(final$vat_l, final$k5, median)

ordered_levels <- names(sort(median_vat))

final$k5 <- factor(final$k5,
                   levels = ordered_levels,
                   labels = seq_along(ordered_levels))

dt_fat <- merge(dt_fat,
                final[, c("ID_751", "sex", "age", "k5")],
                by = c("ID_751", "sex", "age"),
                all.x = TRUE)
rm(final)

############################################################
# Sensitivity analysis: sex-stratified clustering
############################################################

# --------------------
# MEN
# --------------------

fat <- dt_fat %>%
  select("ID_751", "age", "sex",
         "bmat_l1_std", "bmat_l2_std", "vat_l_std", "sat_l_std",
         "heartfat2_std", "ren_hil_ff_std", "liver_pdff_std",
         "smff_std", "imat_std", "pff_std")

fat <- fat[fat$sex == 0, ]
fat <- fat[complete.cases(fat), ]

set.seed(17)
k5 <- kmeans(fat[, -(1:3)], centers = 5, nstart = 25)

fat$k5_sex <- k5$cluster
```

```r
finalm <- merge(dt_fat,
               fat[, c("ID_751", "sex", "age", "k5_sex")],
               by = c("ID_751", "sex", "age"))

finalm$k5_sex <- factor(finalm$k5_sex)
rm(fat)

# --------------------
# WOMEN
# --------------------

fat <- dt_fat %>%
  select("ID_751", "age", "sex",
         "bmat_l1_std", "bmat_l2_std", "vat_l_std", "sat_l_std",
         "heartfat2_std", "ren_hil_ff_std", "liver_pdff_std",
         "smff_std", "imat_std", "pff_std")

fat <- fat[fat$sex == 1, ]
fat <- fat[complete.cases(fat), ]

set.seed(17)
k5 <- kmeans(fat[, -(1:3)], centers = 5, nstart = 25)

fat$k5_sex <- k5$cluster

finalw <- merge(dt_fat,
               fat[, c("ID_751", "sex", "age", "k5_sex")],
               by = c("ID_751", "sex", "age"))

finalw$k5_sex <- factor(finalw$k5_sex)
dt_fat <- rbind(finalm, finalw)
rm(fat, finalw, finalm)

############################################################
# dt_fat now contains in particular:
# 1) k5     = main clustering (sex-combined k-means, k = 5)
# 2) k5_sex = sensitivity analysis (sex-stratified k-means, k = 5)
############################################################

############################################################
# Stability analysis (Supplement Table 3)
# ONLY for main clustering (k5, sex-combined)
# Jaccard similarity via bootstrap resampling (B = 1000)
############################################################

library(fpc)

fat <- dt_fat %>%
  select("bmat_l1_std", "bmat_l2_std", "vat_l_std", "sat_l_std",
         "heartfat2_std", "ren_hil_ff_std", "liver_pdff_std",
         "smff_std", "imat_std", "pff_std")

fat <- fat[complete.cases(fat), ]
```

```r
set.seed(17)
boothc <- clusterboot(data = fat, B = 1000, clustermethod = kmeansCBI,
  k = 5, count = FALSE)
fat$clusterboot <- boothc[["result"]][["result"]][["cluster"]]

# IMPORTANT: clusterboot labels are arbitrary and may differ from previous k5 labels
# visually check VAT distribution across cluster
ggplot(fat, aes(x=as.factor(clusterboot), y= vat_l_std))+ geom_boxplot()

# generate stability table
  stability_table <- data.frame(
    avg_jaccard = round(boothc$bootmean,2),
    instability = boothc$bootbrd)

# Bootstrap cluster labels aligned to the VAT-ordered k5 solution based on
# correspondence of cluster-specific adipose tissue profiles: 1=I, 2=III, 3=II, 4=V,5=IV
stability_table

############################################################
# Radar charts
# Function to generate radar charts as main visualization of clusters
############################################################

library(fmsb)
radar_z <- function(data = dt, cluster_col = "k5",
                    cols = c("bmat_l2_std", "vat_l_std", "sat_l_std", "heartfat2_std",
                             "ren_hil_ff_std", "liver_pdff_std", "smff_std",
                             "imat_std", "pff_std", "bmat_l1_std"),
                    name = c("BMFF L2", "VAT", "SAT", "CFF",
                             "RFF", "HFF", "SMFF", "IMAT", "PFF", "BMFF L1"),
                    cluster_colors = c("#555555", "#66C2A5", "#238B45", "#FDAE61", "#D73027"),
                    sd_range = 2, panel_label = "A") {

  roman <- c("I", "II", "III", "IV", "V")

  layout_matrix <- matrix(c(1, 2, 3,
                            1, 4, 5), nrow = 2, byrow = TRUE)

  layout(layout_matrix, widths = c(1, 1, 1), heights = c(1, 1))
  par(mar = rep(4, 4))

  for (k in 1:5) {

    cl <- data[data[[cluster_col]] == k, cols]
    cl_mean <- apply(cl, 2, mean, na.rm = TRUE)

    # radar input
    df <- rbind(
      rep(sd_range, length(cols)),
      rep(-sd_range, length(cols)),
      cl_mean
    )
```

```r
    colnames(df) <- name
    df <- data.frame(df)

    radarchart(df,
               axistype = 1,
               pcol = cluster_colors[k],
               pfcol = adjustcolor(cluster_colors[k], alpha.f = 0.3),
               cglcol = "grey80",
               cglty = 1,
               plty = 1,
               cglwd = 1,
               plwd = 2,
               vlcex = 1.7,
               axislabcol = "black",
               caxislabels = seq(-sd_range, sd_range, 1))

    titles <- c(
      "I Lean",
      "II Average adiposity",
      "III Bone and muscle adiposity",
      "IV Hepato-abdominal adiposity",
      "V General and pancreatic adiposity"
    )

    label_line <- ifelse(k == 1, -13, 1)

    mtext(titles[k], side = 3, line = label_line, cex = 1.5, font = 2)

    if (k == 1) {
      mtext(panel_label, side = 3, adj = 0, line = 1, cex = 2, font = 2)
    }
  }
}

# Figure 1
radar_z(dt_fat, cluster_col = "k5")

# Suppl figure 6
# women
radar_z(dt_fat[dt_fat$sex==1,], cluster_col = "k5", panel_label = "A")
radar_z(dt_fat[dt_fat$sex==1,], cluster_col = "k5_sex",panel_label = "B")
# men
radar_z(dt_fat[dt_fat$sex==0,], cluster_col = "k5", panel_label = "C")
radar_z(dt_fat[dt_fat$sex==0,], cluster_col = "k5_sex", , panel_label = "D")

# dt2 is data frame containing UK Biobank data only
# dt2 includes participant ID, age, sex, and adipose tissue (AT) variables
# Now, present code for UKB analyses

############################################################
# Define adipose tissue variables used for clustering
############################################################
```

```r
# variable names are aligned with NAKO
AT <- c("vat_l", "sat_l", "liver_pdff", "bmat_l2", "bmat_l1",
        "smff", "imat", "pff", "ren_hil_ff", "heartfat2")

############################################################
# Sex-specific standardisation UKB internally
############################################################

# males
dtm <- dt2[dt2$sex==0,]
m_scaled <- as.data.frame(scale(dtm[, AT]))
colnames(m_scaled) <- str_c(colnames(m_scaled), "_std")
dtm <- cbind(dtm, m_scaled)

# females
dtf <- dt2[dt2$sex==1,]
f_scaled <- as.data.frame(scale(dtf[, AT]))
colnames(f_scaled) <- str_c(colnames(f_scaled), "_std")
dtf <- cbind(dtf, f_scaled)

# combine
dt_fat2 <- rbind(dtf, dtm)
rm(f_scaled, m_scaled, dtf, dtm)

############################################################
# Sex-specific standardisation UKB based on NAKO
############################################################
# get standardization from NAKO
dt_f <- dt[dt$sex == 1, ]
dtN_m <- dt[dt$sex == 0, ]
N_f_scale <- scale(dtN_f[, AT])
N_m_scale <- scale(dtN_m[, AT])

# standradize UKB sex-specifically based on NAKO
dt2_f <- dt_fat2[dt_fat2$sex == 1, ]
dt2_m <- dt_fat2[dt_fat2$sex == 0, ]

dt2_f_scaled <- as.data.frame(
  scale(dt2_f[, names(AT)],
        center = attr(N_f_scale, "scaled:center"),
        scale  = attr(N_f_scale, "scaled:scale")))
colnames(dt2_f_scaled) <- paste0(names(col_map), "_std_N")

dt2_m_scaled <- as.data.frame(
  scale(dt2_m[, names(AT)],
        center = attr(N_m_scale, "scaled:center"),
        scale  = attr(N_m_scale, "scaled:scale")))
colnames(dt2_m_scaled) <- paste0(names(AT), "_std_N")

dt2_f <- cbind(dt2_f, dt2_f_scaled)
dt2_m <- cbind(dt2_m, dt2_m_scaled)
```

```r
# combine
dt_fat2 <- rbind(dt2_f, dt2_m)

############################################################
# dt_fat2 now contains UKB data in particular:
# 1) UKB internally standardized AT, e.g vat_l_std
# 2) UKB relative to NAKO standardized AT, e.g. vat_l_std_N
############################################################

############################################################
# UKB clustering based on internally standardized AT
############################################################

# select same feature set as in NAKO clustering
new_fat <- dt_fat2 %>%
  select(ID, age, sex, bmat_l1_std, bmat_l2_std, vat_l_std, sat_l_std,
         heartfat2_std, ren_hil_ff_std, liver_pdff_std, smff_std, imat_std, pff_std)

# complete-case analysis
new_fat_complete <- new_fat[complete.cases(new_fat), ]

# assign each subject to nearest centroid (Euclidean distance)
new_clusters <- apply(new_fat_complete[, -(1:3)] , 1,
                     function(x) {which.min(colSums((t(centroids) - x)^2))})
new_fat_complete$k5 <- new_clusters

# merge cluster assignment back to full UKB dataset
dt_fat2 <- dt %>% left_join(new_fat_complete %>% select(ID, k5), by = c("ID"))

# align cluster labels by median VAT (for interpretability)
median_vat <- tapply(dt_fat2$vat_l, dt_fat2$k5, median)
ordered_levels <- names(sort(median_vat))
dt_fat2$k5 <- factor(dt_fat2$k5, levels = ordered_levels, labels = seq_along(ordered_levels))
rm(new_fat, new_clusters, new_fat_complete)

############################################################
# UKB cluster assignment using NAKO-standardized UKB variables
############################################################
# select UKB variables standardized using NAKO parameters
new_fat <- dt %>%
  select(ID, age, sex,
         bmat_l1_std_N, bmat_l2_std_N, vat_l_std_N, sat_l_std_N,
         heartfat2_std_N, ren_hil_ff_std_N, liver_pdff_std_N,
         smff_std_N, imat_std_N, pff_std_N)

# harmonize variable names to match centroid space

new_fat <- new_fat %>%
  rename(
    heartfat2_std = heartfat2_std_N,
    bmat_l1_std   = bmat_l1_std_N,
    bmat_l2_std   = bmat_l2_std_N,
    vat_l_std     = vat_l_std_N,
```

```
    sat_l_std     = sat_l_std_N,
    ren_hil_ff_std = ren_hil_ff_std_N,
    liver_pdff_std = liver_pdff_std_N,
    smff_std       = smff_std_N,
    imat_std       = imat_std_N,
    pff_std        = pff_std_N)

# complete-case analysis
new_fat_complete <- new_fat[complete.cases(new_fat), ]

# assign each subject to nearest centroid (Euclidean distance)
new_clusters <- apply(new_fat_complete[, -(1:3)], 1,
                     function(x) {which.min(colSums((t(centroids) - x)^2))})
new_fat_complete$k5N <- new_clusters

# merge cluster assignment back to full UKB dataset
dt_fat2 <- dt_fat2 %>% left_join(new_fat_complete %>% select(ID, k5N), by = "ID")

# align cluster labels by median VAT (for interpretability)
median_vat <- tapply(dt_fat2$vat_l, dt_fat2$k5N, median)
ordered_levels <- names(sort(median_vat))
dt_fat2$k5N <- factor(dt_fat2$k5N, levels = ordered_levels, labels = seq_along(ordered_levels))

############################################################
# dt_fat2 now contains UKB data in particular:
# 1) k5 as kmeans k = 5 clustering as defined by assigning UKB data
# with internal standardization to NAKO centroids
# 2) k5N as kmeans k = 5 clustering as defined by assigning UKB data
# with NAKO-based standardization to NAKO centroids
############################################################

############################################################
#Create visualizations to assess robustness of cluster validation
############################################################

##########
# Figure 1
radar_z(dt_final2, cluster_col = "k5", panel_label = "B")
##########

#############################
# Suppl  Figure 1 Pie charts
#############################

bar_fill <- c("1" = "#555555",
              "2" = "#66C2A5",
              "3" = "#238B45",
              "4" = "#FDAE61",
              "5" = "#D73027")

# summarize UKB data based on k5 clustering
dt_UK_summary <- dt_fat2 %>%
```

```
  filter(!is.na(k5)) %>%
  count(k5) %>%
  mutate(pct = n / sum(n) * 100,
         label = paste0(n, "\n", round(pct, 1), "%"))

# generate pie
p1_pie <- ggplot(dt_UK_summary, aes(x = 1, y = pct, fill = factor(k5))) +
  geom_bar(stat = "identity", color = "white", width = 1) +
  coord_polar(theta = "y") +
  geom_text(aes(label = label), size = 4, position = position_stack(vjust = 0.5)) +
  scale_fill_manual(values = bar_fill, labels = cluster_labels) +
  labs(title = "UKB", subtitle = "internal standardization", fill = "Cluster") +
  theme_void() +
  theme(legend.position = "bottom",
        plot.title = element_text(size = 16, face = "bold", hjust = 0.5),
        legend.title = element_text(size = 15), legend.text = element_text(size = 15),
        plot.subtitle = element_text(size = 15, hjust = 0.5))

# summarize NAKO data based on main clustering
dt_NAKO_summary <- dt %>%
  filter(!is.na(k5)) %>%
  count(k5) %>%
  mutate(pct = n / sum(n) * 100,
         label = paste0(n, "\n", round(pct, 1), "%"))

# generate pie
p2_pie <- ggplot(dt_NAKO_summary, aes(x = 1, y = pct, fill = factor(k5))) +
  geom_bar(stat = "identity", color = "white", width = 1) +
  coord_polar(theta = "y") +
  geom_text(aes(label = label), size = 4, position = position_stack(vjust = 0.5)) +
  scale_fill_manual(values = bar_fill, labels = cluster_labels) +
  labs(title = "NAKO", fill = "Cluster") +
  theme_void() +
  theme(legend.position = "none",
        plot.title = element_text(size = 16, face = "bold", hjust = 0.5))

# summarize UKB data based on k5N clustering
dt_UK_summary2 <- dt_fat2 %>% count(k5N) %>%
  mutate(pct = n / sum(n) * 100,
         label = paste0(n, "\n", round(pct, 1), "%"))

# generate pie
p3_pie <- ggplot(dt_UK_summary2, aes(x = 1, y = pct, fill = factor(k5N))) +
  geom_bar(stat = "identity", color = "white", width = 1) +
  coord_polar(theta = "y") +
  geom_text(aes(label = label), size =4, position = position_stack(vjust = 0.5)) +
  scale_fill_manual(values = bar_fill, labels = cluster_labels) +
  labs(title = "UKB ",subtitle = "NAKO-based standardization",  fill = "Cluster") +
  theme_void() +
  theme(legend.position = "none",
        plot.title = element_text(size = 16, face = "bold", hjust = 0.5),
        plot.subtitle = element_text(size = 15, hjust = 0.5))
```

```
# combine pie charts
p2_pie | p1_pie | p3_pie

########################################
# Suppl figure 3 PCA plots NAKO and UKB
########################################

# get variable names
cols <- c("bmat_l1_std", "bmat_l2_std", "vat_l_std", "sat_l_std", "heartfat2_std",
          "ren_hil_ff_std", "liver_pdff_std", "smff_std", "imat_std", "pff_std")

colsN <- c("bmat_l1_std_N", "bmat_l2_std_N", "vat_l_std_N", "sat_l_std_N", "heartfat2_std_N",
           "ren_hil_ff_std_N", "liver_pdff_std_N", "smff_std_N", "imat_std_N", "pff_std_N")

# NAKO
pca <- prcomp(dt_fat[,cols])
pca_data <- as.data.frame(pca$x)
pca_data$k5 <- as.factor(dt_fat$k5)

(pca$sdev^2 / sum(pca$sdev^2)*100)[1:2] #  % of explained variance of first 2 components

ggplot(pca_data, aes(x = PC1, y = PC2, color = k5, fill = k5)) +
  geom_point(alpha = 0.1, size = 2) +
  stat_ellipse(geom = "polygon", alpha = 0.2, color = NA) +
  stat_ellipse(geom = "path", linewidth = 1.2) +
  scale_color_manual(values = cluster_colors, labels = cluster_labels, name = "Cluster") +
  scale_fill_manual(values = cluster_colors, labels = cluster_labels, name = "Cluster") +
  xlim(-8,8) +
  labs(x = "PC1 (53.5%)", y = "PC2 (12.0%)") +
  my_theme + ggtitle("NAKO")

# UKB with clustering internally standardization based
pca <- prcomp(dt_fat2[,cols])
pca_data <- as.data.frame(pca$x)
pca_data$k5 <- as.factor(dt_fat2$k5)

(pca$sdev^2 / sum(pca$sdev^2)*100)[1:2] #  % of explained variance of first 2 components

ggplot(pca_data, aes(x = PC1, y = PC2, color = k5, fill = k5)) +
  geom_point(alpha = 0.1, size = 2) +
  stat_ellipse(geom = "polygon", alpha = 0.2, color = NA) +
  stat_ellipse(geom = "path", linewidth = 1.2) +
  scale_color_manual(values = cluster_colors, labels = cluster_labels, name = "Cluster") +
  scale_fill_manual(values = cluster_colors, labels = cluster_labels, name = "Cluster") +
  xlim(-8,8) +
  labs(x = "PC1 (44.5%)", y = "PC2 (17.2%)", title = "UKB", subtitle = "internal standardization") +
  my_theme

# UKB with clustering NAKO standardization based
pca <- prcomp(dt_fat2[,colsN])
pca_data <- as.data.frame(pca$x)
pca_data$k5N <- as.factor(dt_fat2$k5N)
```

```r
(pca$sdev^2 / sum(pca$sdev^2)*100)[1:2] # % of explained variance of first 2 components

ggplot(pca_data, aes(x = PC1, y = PC2, color = k5N, fill = k5N)) +
  geom_point(alpha = 0.1, size = 2) +
  stat_ellipse(geom = "polygon", alpha = 0.2, color = NA) +
  stat_ellipse(geom = "path", linewidth = 1.2) +
  scale_color_manual(values = cluster_colors, labels = cluster_labels, name = "Cluster") +
  scale_fill_manual(values = cluster_colors, labels = cluster_labels, name = "Cluster") +
  xlim(-8,8) +
  labs(x = "PC1 (44.8%)", y = "PC2 (16.3%)", title = "UKB", subtitle = "NAKO-based standardization") +
  my_theme

###################################
# Suppl figure 4 silhouette plots
###################################

# compute silhouette widths for NAKO
dfat <- dist(dt_fat[,cols])
clusters <- as.numeric(dt_fat$k5)
sil = cluster::silhouette(clusters, dfat)

# NAKO silhouette plot
fviz_silhouette(sil)  + labs(title = "NAKO") +
  scale_color_manual(values = cluster_colors) +
  scale_fill_manual(values = cluster_colors) +
  scale_y_continuous(limits = c(-0.25,1), breaks = seq(-0.25,1,0.25))+
  my_theme + ylab("Silhouette width") + theme(axis.text.x=element_blank(),axis.ticks.x=element_blank())
rm(sil)

# compute silhouette widths for UKB internally standardized clustering
dfat <- dist(dt_fat2[,cols])
clusters <- as.numeric(dt_fat2$k5)
sil = cluster::silhouette(clusters, dfat)

fviz_silhouette(sil)  + labs(title = "UKB", subtitle = "internal standardization") +
  scale_color_manual(values = cluster_colors) +
  scale_fill_manual(values = cluster_colors) +
  scale_y_continuous(limits = c(-0.25,1), breaks = seq(-0.25,1,0.25))+
  my_theme + ylab("Silhouette width") + theme(axis.text.x=element_blank(),axis.ticks.x=element_blank())
rm(sil)

# compute silhouette widths for UKB NAKO-based standardized clustering
dfatk <- dist(dt_fat2[,colsN])
clustersk <- as.numeric(dt_fat2$k5N)
sil2 = cluster::silhouette(clustersk, dfatk)

fviz_silhouette(sil2)  + labs(title = "UKB", subtitle = "NAKO-based standardization") +
  scale_color_manual(values = cluster_colors) +
  scale_fill_manual(values = cluster_colors) +
  scale_y_continuous(limits = c(-0.25,1), breaks = seq(-0.25,1,0.25))+
  my_theme + ylab("Silhouette width") + theme(axis.text.x=element_blank(),axis.ticks.x=element_blank())
rm(sil2)
```

```
####################################
# Suppl figure 5: forest plots
####################################

# results based on KORA data were derived in previous
# publication https://doi.org/10.1016/j.metabol.2024.156130
# linear regression models for risk scores and main clustering in NAKO
# variables myscore2, FRS10A and FRS30A contain the risk scores as
# calculated by the respective equations
# references for the equations are found in supplementary text 2
# risk scores are log transformed due to skewed distribution and
# models are unadjusted since the risk scores incorporate particularly age, sex
score2 <- lm(log(myscore2) ~ k5, data = dt_fat)
frs10 <- lm(log(FRS10A) ~ k5, data = dt_fat)
frs30 <- lm(log(FRS30A) ~ k5, data = dt_fat)

# helper function
extract_results <- function(model, dataset_label) {
  data.table(
    dataset = dataset_label,
    exposure = factor(c("II","III","IV","V")),
    beta = exp(coef(model)[2:5]),
    lowerCI = exp(confint(model)[2:5, 1]),
    upperCI = exp(confint(model)[2:5, 2]))}

#
# Extract regression results for SCORE2
dat_score2 <- rbind(extract_results(score2, "NAKO"))

# Extract regression results for FRS10
dat_frs10 <- rbind(extract_results(frs10, "NAKO"))

# Extract regression results for FRS30
dat_frs30 <- rbind(extract_results(frs30, "NAKO"))

# Plot for SCORE2
ggplot(dat_score2, aes(x=exposure, y=beta,color=exposure) +
  geom_errorbar(aes(ymin=lowerCI, ymax=upperCI),
                width=0.4, position=position_dodge(width=0.5)) +
  geom_point(size=3.5, position=position_dodge(width=0.5)) +
  scale_color_manual(values=color_code, name="Cluster") +  # legend for colors
  labs(x=NULL,
       y="Relative increase (exp(ß); 95% CI)\n in CVD risk compared to cluster I") +
  geom_hline(yintercept=1, lty=2, color="grey20", size=0.3) +
  coord_cartesian(clip = "off") +
  scale_y_continuous(breaks = seq(0, 5, by=1), limits = c(0, 6)) +
  my_theme +
  theme(legend.position = "none", axis.text.x = element_blank(),
        axis.title.y = element_text(size =15, margin = margin(r = 25))) +
  ggtitle("SCORE2")

# Plot for FRS10
ggplot(dat_frs10, aes(x=exposure, y=beta, color=exposure)) +
```

```r
  geom_errorbar(aes(ymin=lowerCI, ymax=upperCI),
                width=0.4, position=position_dodge(width=0.5)) +
  geom_point(size=3.5, position=position_dodge(width=0.5)) +
  scale_color_manual(values=color_code, name="Cluster") +
  labs(x=NULL, y=NULL) +
  geom_hline(yintercept=1, lty=2, color="grey20", size=0.3) +
  coord_cartesian(clip = "off") +
  scale_y_continuous(breaks = seq(0, 5, by=1), limits = c(0, 6)) +
  my_theme +
  theme(legend.position = "none", axis.title.y = element_blank(),
        axis.text.y = element_blank(),axis.text.x = element_blank(),
        axis.ticks.y = element_blank()) +
  ggtitle("FRS10")

# Plot for FRS30
plot3 <- ggplot(dat_frs30, aes(x=exposure, y=beta, color=exposure)) +
  geom_errorbar(aes(ymin=lowerCI, ymax=upperCI),
                width=0.4, position=position_dodge(width=0.5)) +
  geom_point(size=3.5, position=position_dodge(width=0.5)) +
  scale_color_manual(values=color_code, name="Cluster") +
  labs(x=NULL, y=NULL) +
  geom_hline(yintercept=1, lty=2, color="grey20", size=0.3) +
  coord_cartesian(clip = "off") +
  scale_y_continuous(breaks = seq(0, 5, by=1), limits = c(0, 6)) +
  my_theme +
  theme(axis.title.y = element_blank(),axis.text.x = element_blank(),
        axis.text.y = element_blank(),
        axis.ticks.y = element_blank()) +
  ggtitle("FRS30")

#######################################################
# Suppl figure 7: forest plots sex-stratified models
#######################################################

# linear models sex-stratified analyses and main clustering
score2f <- lm(log(myscore2) ~ k5, data = dt_fat[dt_fat$sex==1,])
score2m <- lm(log(myscore2) ~ k5, data = dt_fat[dt_fat$sex==0,])
frs10f <- lm(log(FRS10A) ~ k5, data = dt_fat[dt_fat$sex==1,])
frs10m <- lm( log(FRS10A)~ k5, data = dt_fat[dt_fat$sex==0,])
frs30f <- lm(log(FRS30A) ~ k5, data = dt_fat[dt_fat$sex==1,])
frs30m <- lm(log(FRS30A) ~ k5, data = dt_fat[dt_fat$sex==0,])

# extract results
dat_score2 <- rbind(
  extract_results(score2f, "Female"),
  extract_results(score2m, "Male"))

dat_frs10 <- rbind(
  extract_results(frs10f, "Female"),
  extract_results(frs10m, "Male"))

dat_frs30 <- rbind(
  extract_results(frs30f, "Female"),
```

```r
  extract_results(frs30m, "Male"))

# plot SCORE2 main clustering
ggplot(dat_score2, aes(x=exposure, y=beta,
                                color=exposure, shape=dataset)) +
  geom_errorbar(aes(ymin=lowerCI, ymax=upperCI),
              width=0.4, position=position_dodge(width=0.5)) +
  geom_point(size=3.5, position=position_dodge(width=0.5)) +
  scale_color_manual(values=color_code, name="Cluster") +  # legend for colors
  scale_shape_manual(values=c(16, 17), name="Sex") +   # legend for shapes
  labs(x=NULL,
       y="Relative increase (exp(ß); 95% CI)\n in CVD risk compared to cluster I") +
  geom_hline(yintercept=1, lty=2, color="grey20", size=0.3) +
  coord_cartesian(clip = "off") +
  scale_y_continuous(breaks = seq(0, 5, by=1), limits = c(0, 6)) +
  my_theme +
  theme(legend.position = "none", axis.text.x = element_blank(),
        axis.title.y = element_text(size =15, margin = margin(r = 25))) +
  ggtitle("SCORE2")

# plot FRS10 main clustering
ggplot(dat_frs10, aes(x=exposure, y=beta,
                               color=exposure, shape=dataset)) +
  geom_errorbar(aes(ymin=lowerCI, ymax=upperCI),
              width=0.4, position=position_dodge(width=0.5)) +
  geom_point(size=3.5, position=position_dodge(width=0.5)) +
  scale_color_manual(values=color_code, name="Cluster") +
  scale_shape_manual(values=c(16, 17), name="Sex") +
  labs(x=NULL, y=NULL) +
  geom_hline(yintercept=1, lty=2, color="grey20", size=0.3) +
  coord_cartesian(clip = "off") +
  scale_y_continuous(breaks = seq(0, 5, by=1), limits = c(0, 6)) +
  my_theme +
  theme(legend.position = "none", axis.title.y = element_blank(),
        axis.text.x = element_blank()) +
  ggtitle("FRS10")

# plot FRS30 main clustering
ggplot(dat_frs30, aes(x=exposure, y=beta,
                               color=exposure, shape=dataset)) +
  geom_errorbar(aes(ymin=lowerCI, ymax=upperCI),
              width=0.4, position=position_dodge(width=0.5)) +
  geom_point(size=3.5, position=position_dodge(width=0.5)) +
  scale_color_manual(values=color_code, name="Cluster") +
  scale_shape_manual(values=c(16, 17), name="Sex") +
  labs(x=NULL, y=NULL) +
  geom_hline(yintercept=1, lty=2, color="grey20", size=0.3) +
  coord_cartesian(clip = "off") +
  scale_y_continuous(breaks = seq(0, 5, by=1), limits = c(0, 6)) +
  my_theme +
  theme(axis.title.y = element_blank(),axis.text.x = element_blank()) +
  ggtitle("FRS30")
```

```r
rm(score2f, score2m, frs10f, frs10m, frs30f, frs30m)

# linear models sex-stratified analyses and sex-specific clustering
score2f <- lm(log(myscore2) ~ k5_sex, data = dt_fat[dt_fat$sex==1,])
score2m <- lm(log(myscore2) ~ k5_sex, data = dt_fat[dt_fat$sex==0,])
frs10f <- lm(log(FRS10A) ~ k5_sex, data = dt_fat[dt_fat$sex==1,])
frs10m <- lm( log(FRS10A)~ k5_sex, data = dt_fat[dt_fat$sex==0,])
frs30f <- lm(log(FRS30A) ~ k5_sex, data = dt_fat[dt_fat$sex==1,])
frs30m <- lm(log(FRS30A) ~ k5_sex, data = dt_fat[dt_fat$sex==0,])

# extract and plot as before

######################################################
# Suppl figure 8: Subgroup analyses by BMI and waist circumference (WC)
######################################################

# take cutoffs from doi.org/10.1038/s41574-019-0310-7
# men with elevated BMI and WC
dt1 <- dt_fat[which(dt_fat$sex==0 & dt_fat$waist>=100 & (dt_fat$bmi>=25 & dt_fat$bmi<30)),]
table(dt1$k5)
radar_z(dt1, cluster_col = "k5", panel_label = "B")

# women with elevated BMI and WC
dt2 <- dt_fat[which(dt_fat$sex==1 & (dt_fat$bmi>=25 & dt_fat$bmi<30) & dt_fat$waist>=90),]
table(dt2$k5)
radar_z(dt2, cluster_col = "k5", panel_label = "A")
```
